# Accuracy of computer-aided chest X-ray screening in the Kenya National Tuberculosis Prevalence Survey

**DOI:** 10.1101/2021.10.21.21265321

**Authors:** Brenda Mungai, Jane Ong‘angò, Chu Chang Ku, Marc YR Henrion, Ben Morton, Elizabeth Joekes, Elizabeth Onyango, Richard Kiplimo, Dickson Kirathe, Enos Masini, Joseph Sitienei, Veronica Manduku, Beatrice Mugi, The IMPALA Consortium, Stephen Bertel Squire, Peter MacPherson

## Abstract

**Background:** Community-based screening for tuberculosis (TB) could improve detection but is resource intensive. We set out to evaluate the accuracy of computer-aided TB screening using digital chest X-ray (CXR) to determine if this approach met target product profiles (TPP) for community-based screening.

**Methods:** CXR images from participants in the 2016 Kenya National TB Prevalence Survey were evaluated using CAD4TBv6 (Delft Imaging), giving a probabilistic score for pulmonary TB ranging from 0 (low probability) to 99 (high probability). We constructed a Bayesian latent class model to estimate the accuracy of CAD4TBv6 screening compared to bacteriologically-confirmed TB across CAD4TBv6 threshold cut-offs, incorporating data on Clinical Officer CXR interpretation, participant demographics (age, sex, TB symptoms, previous TB history), and sputum results. We compared model-estimated sensitivity and specificity of CAD4TBv6 to optimum and minimum TPPs.

**Results:** Of 63,050 prevalence survey participants, 61,848 (98%) had analysable CXR images, and 8,966 (14.5%) underwent sputum bacteriological testing; 298 had bacteriologically-confirmed pulmonary TB. Median CAD4TBv6 scores for participants with bacteriologically-confirmed TB were significantly higher (72, IQR: 58-82.75) compared to participants with bacteriologically-negative sputum results (49, IQR: 44-57, p<0.0001). CAD4TBv6 met the optimum TPP; with the threshold set to achieve a mean sensitivity of 95% (optimum TPP), specificity was 83.3%, (95% credible interval [CrI]: 83.0%—83.7%, CAD4TBv6 threshold: 55). There was considerable variation in accuracy by participant characteristics, with older individuals and those with previous TB having lowest specificity.

**Conclusions:** CAD4TBv6 met the optimal TPP for TB community screening. To optimise screening accuracy and efficiency of confirmatory sputum testing, we recommend that an adaptive approach to threshold setting is adopted based on participant characteristics.

**Take home message:** CAD4TBv6 met the optimal WHO target product profile for a community TB screening tool. Specificity was lower in adults with previous TB and those aged 41 years or older; an adaptive approach to setting CAD thresholds will likely be required to optimize use.

## Introduction

With over 95% tuberculosis (TB) cases and deaths occurring in developing countries, there is need for substantially improved case detection to find the “missing millions” and accelerate action to achieve the sustainable development goals to end TB by 2030. (1-4) Chest radiography (CXR) with computer-aided detection (CAD) software for TB has been recommended for systematic screening for tuberculosis disease in the most recent WHO TB Screening Guidelines. (5) However, supporting data have predominantly come from clinical settings and CAD diagnostic accuracy is likely to vary considerably across different screening strategies and populations. (6-8)

CXRs were used extensively in TB screening and active case finding (ACF) programmes in the mid-20th century due to high sensitivity (94%, 95% CI 88–98%), potential for high throughput, and lower infectious risk to health workers (compared to sputum collection for all). (9-12) In addition, CXR can detect infectious but asymptomatic TB patients, this is important as a substantial fraction of TB transmission is attributable to the often prolonged asymptomatic infectious period.(13) Barriers to widespread CXR use include limited access to high quality radiography equipment, critical shortage of radiologists in low- and middle-income countries (LMICs), and inter-and intra-observer variations during interpretation. (11, 12, 14) CAD software that provides a probabilistic score for TB offers a potential solution to these limitations.(6, 7, 15, 16) Previous evaluations of CAD software have been mostly conducted in triage testing use situations, with very little data available to evaluate accuracy in community-based TB screening interventions.(6, 8, 16, 17)

Our aim was to evaluate the accuracy of the Computer-Aided Detection for Tuberculosis version 6 (CAD4TBv6) system for TB screening using a large data set (n=61,848) from the 2016 Kenya National TB prevalence survey.(18, 19) To do this we used a Bayesian modelling approach to evaluate the accuracy of CAD4TBv6 and Clinical Officer CXR interpretation against the bacteriological reference standard used within the prevalence survey. We hypothesized that CAD4TBv6 diagnostic sensitivity and specificity would meet the target product profile (TPP) for a test to identify people suspected of having TB, but that accuracy would vary between population groups, implying that an adaptive approach to CAD screening would be required.(20)

## Methods

### Study design

We conducted a retrospective analysis of cross-sectional individual-level participant data from adult community members who participated in the 2016 Kenya National TB Prevalence Survey.(19)

### Study population and Kenya TB prevalence survey procedures

The 2016 Kenya prevalence survey was undertaken to determine the prevalence of bacteriologically confirmed pulmonary TB among adults aged 15 years and older and used the WHO-recommended screening strategy including symptom questionnaire and CXR.(21) It has been reported fully elsewhere.(18, 19) The survey found a weighted national pulmonary TB prevalence of 558 (95% CI 455-662) per 100,000 adult population.(18, 19)

Prevalence survey participants completed a questionnaire to elicit TB symptoms. Subsequently, a digital posterior anterior CXR (CXDI, Delft Imaging, The Netherlands) was digitally acquired and uploaded to a digital archive. Study Clinical Officers who had received intensive one-week training in CXR interpretation independently read each film, blinded to bacteriological results (Clinical Officer characteristic are summarised in Supplementary Text 1). Clinical Officers classified CXRs as either: A) normal; B) abnormal, suggestive of TB; or C) abnormal other, in line with published definitions.(19) Participants with a CXR classified as “abnormal, suggestive of TB” by either one of the Clinical Officers, or with a cough of more than two weeks or who declined CXR screening were eligible for sputum collection.

Two sputum samples (spot and following morning) were obtained from eligible participants and transported to the National Tuberculosis Reference Laboratory and tested with Xpert MTB/Rif. Solid culture using Lowenstein Jensen medium (incubation at 37°C) was conducted on all samples and reported as negative if there was no growth after eight weeks. To confirm the presence of *Mycobacterium tuberculosis* complex, all visible colonies grown on culture media were confirmed by acid-fast bacilli (AFB) microscopy and tested for presence of Mycobacterium protein 64 (MPT64) by SD Bio line Immunochromatographical assay. Geno-Type Mycobacterium AS (Hain Life Science) test kits were used to identify presence of non-tuberculous mycobacterium.(18, 19)

Participants with bacteriologically-confirmed pulmonary TB (sputum Xpert and/or culture-positive) were referred for TB treatment; participants with CXR abnormalities and no bacteriological evidence of TB were linked to health facilities for clinical assessment. HIV testing was not undertaken as part of TB prevalence survey activities. In line with national guidelines, participants referred for TB treatment were offered HIV testing at referral facilities.

### Study procedures and definitions

Analysis was conducted between January 2020 and October 2021. Anonymised, compressed DICOM CXR images were uploaded from the prevalence survey digital archive to the Delft Imaging CAD4TB cloud server and analysed using CAD4TBv6. (22) Results were provided as probabilistic scores, ranging from 0 to 99, with higher scores indicating a greater probability of TB. The reference standard for this analysis was bacteriologically-confirmed TB, defined as sputum Xpert and/or culture positive with MTB speciation. Analysis was conducted independently; the commercial provider (Delft Imaging) was not part of the study team and had no role in study design, data collection, analysis, or interpretation of results.

### Statistical analysis

The characteristics of prevalence survey participants were summarized using means (with standard deviations), medians (with interquartile ranges), and percentages, and compared by Clinical Officer CXR interpretation. We used the Kruskall-Wallis test to investigate differences in CAD4TBv6 scores between Clinical Officer interpretation groups, and Chi-square and Fisher’s exact tests for categorical participant characteristics. Distributions of CAD4TBv6 scores were summarized by medians and 95% highest density intervals (HDI) and compared by whether sputum was collected or not, and by sputum bacteriological status.

For our primary study outcome, we compared the accuracy (sensitivity, specificity, and area under receiver-operator curve [AUC]) of CAD4TBv6 with the bacteriological reference standard. As collection of sputum was conditional on either a participant reporting having cough of two weeks or greater or a Clinical Officer CXR classification of “abnormal, suggestive of TB”, Bayesian latent class modelling was employed to infer disease prevalence within the portion of the study population without TB symptoms or CXR signs suggestive of TB, and to estimate the sensitivity, specificity, and AUC of CAD4TBv6 at thresholds ranging from 0 to 99. The model also outputs estimates of the underlying status of active pulmonary TB, and the sensitivity and specificity of Clinical Officer CXR interpretation as a screening tool and of sputum bacteriological results for the underlying true TB status. Full model details and diagnostics are reported in Supplemental Text 2. We placed informative priors on the overall prevalence of TB, inferred by the prevalence survey results, and weakly informative priors on other model parameters. To aid model convergence, we fixed specificity of the combined bacteriological reference standard (Xpert or culture positive) to be 99%.

Models were fitted in Stan using the cmdstanr interface, with convergence assessed by inspecting trace plots across three sampling chains and Gelman-Rubin statistics. Inference was based on summarising 12,000 post-warm-up samples. We plotted model posterior summary estimates of sensitivity and specificity across CAD4TBv6 thresholds, and compared to optimum (sensitivity: 95%, specificity: 80%) and minimum (sensitivity: 90%, specificity: 70%) TPP for a community or referral test to identify people suspected of having TB. (20) In secondary analysis, we restricted model sensitivity and specificity estimates for participants by age group, sex, chronic cough status and history of previous TB treatment, and summarised as a function of CAD4TBv6 threshold, estimating what accuracy would be achieved by setting an overall screening CAD4TBv6 threshold to achieve the optimum TPP sensitivity cut-off (95%). We did not stratify by HIV status, as there was substantial missing data and testing was not performed in the prevalence survey. All analysis was done using R v4.1.1 (R Foundation for Statistical Computing, Vienna).

### Ethical considerations

This study was conducted as part of the Kenya Prevalence survey ethics approval reference number SSC 2094 by the Kenya Medical Research Institute. Prevalence survey participants provided written informed consent for survey activities. Additional approval was obtained from the Division of National Tuberculosis, Leprosy and Lung Disease program which is the custodian of the TB prevalence survey data. The data was processed within the Kenya Health Informatics System Governance (23) and General Data Protection Regulations.(24) All CXR images were deleted from the Delft Imaging server after analysis as contractually stipulated.

## Results

### Participant characteristics and Field Reader chest X-ray interpretation

A total of 62,484 CXR images were uploaded for CAD4TBv6 processing. After exclusion of 636 images that were either not analysable by the CAD4TBv6 software or had missing clinical data, 61,848 (99.0%) were included for analysis (Figure 1).

**Figure 1:**
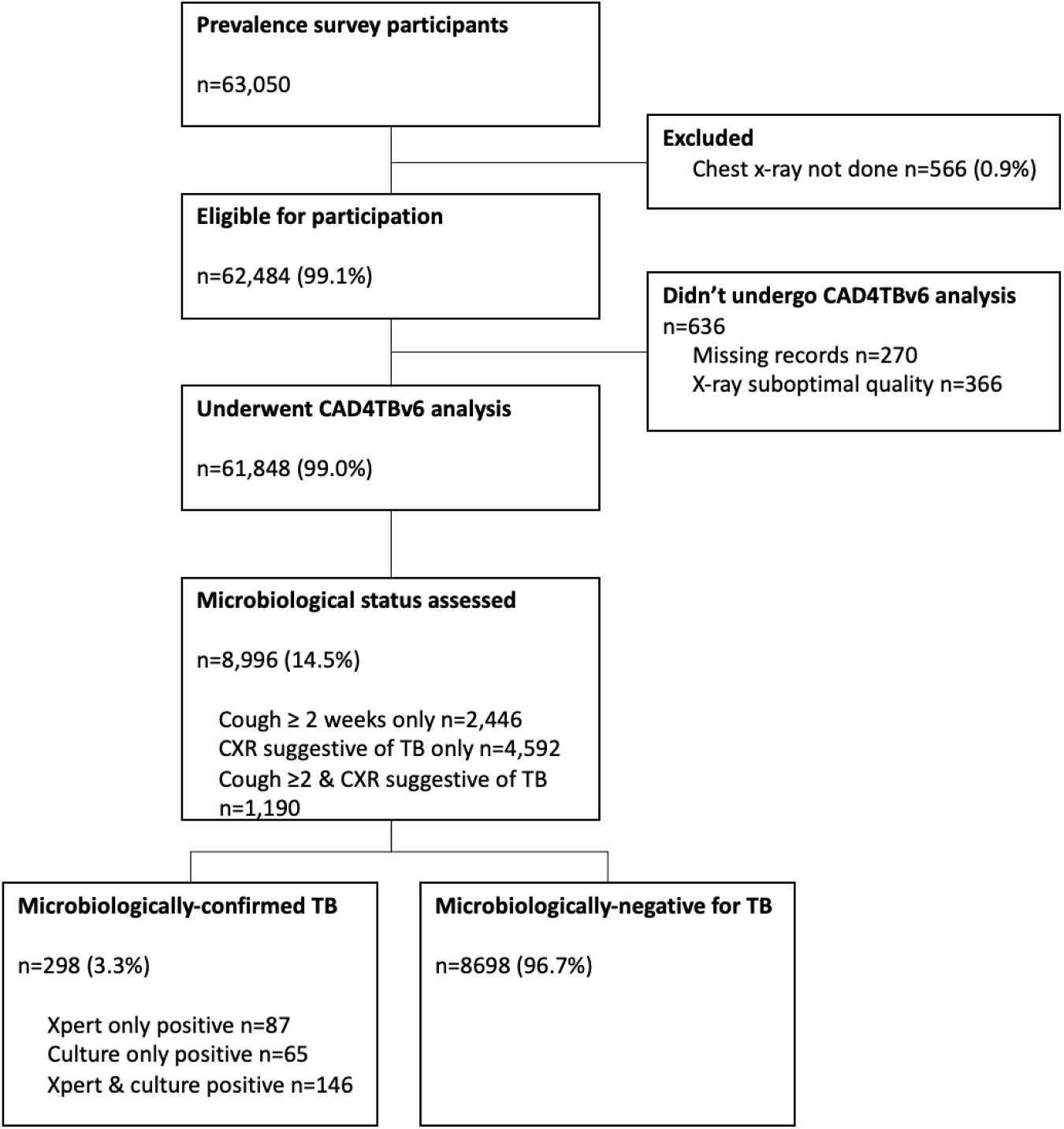
Participant flowchart and results of prevalence survey investigations.

Of the 61,848 participants whose images were analysed, 58.5% (36,187) were women and 70.7% (43,754) were aged <45 years (Table 1). Two thousand and eighty-four (3.4%) had previously been treated for TB, and 58 (0.1%) were currently being treated for TB. Overall, HIV positive status was self-reported by 1,577/31,495 (5.0%) of participants with data available.

**Table 1:** Characteristics of participants in Kenya National TB prevalence survey, by Clinical Officer chest X-ray interpretation.

|  | Normal<br>(N=50,045) | Abnormal, other<br>(N=5,045) | Suggestive of TB<br>(N=6,758) | Total<br>(N=61,848) | P-value<sup>†</sup> |
| --- | --- | --- | --- | --- | --- |
| **Age group** |  |  |  |  | < 0.001 |
| 15 – 24 years | 16,305 (32.6%) | 403 (8.0%) | 742 (11.0%) | 17,450 (28.2%) |  |
| 25 - 34 years | 13,400 (26.8%) | 627 (12.4%) | 1,167 (17.3%) | 15,194 (24.6%) |  |
| 35 - 44 years | 9,072 (18.1%) | 765 (15.2%) | 1,273 (18.8%) | 11,110 (18.0%) |  |
| 45 - 54 years | 5,553 (11.1%) | 831 (16.5%) | 1,032 (15.3%) | 7,416 (12.0%) |  |
| 55 - 64 years | 3,210 (6.4%) | 901 (17.9%) | 957 (14.2%) | 5,068 (8.2%) |  |
| 65+ years | 2,505 (5.0%) | 1,518 (30.1%) | 1,587 (23.5%) | 5,610 (9.1%) |  |
| **Sex** |  |  |  |  | < 0.001 |
| Women | 29,432 (58.8%) | 3,410 (67.6%) | 3,345 (49.5%) | 36,187 (58.5%) |  |
| Men | 20,613 (41.2%) | 1,635 (32.4%) | 3,413 (50.5%) | 25,661 (41.5%) |  |
| **HIV-status\*** |  |  |  |  |  |
| Missing | 23,895 | 2871 | 3,587 | 30,353 |  |
| HIV-negative | 24,985 (95.5%) | 2,055 (94.5%) | 2,878 (90.8%) | 29,918 (95.0%) | < 0.001 |
| HIV-positive | 1,165 (4.5%) | 119 (5.5%) | 293 (9.2%) | 1,577 (5.0%) |  |
| **Cough** |  |  |  |  | < 0.001 |
| No | 43,989 (87.9%) | 4,065 (80.6%) | 4,752 (70.3%) | 52,806 (85.4%) |  |
| Yes | 6,056 (12.1%) | 980 (19.4%) | 2,006 (29.7%) | 9,042 (14.6%) |  |
| **Cough>2 weeks** |  |  |  |  | < 0.001 |
| No | 47,859 (95.6%) | 4,504 (89.3%) | 5,484 (81.1%) | 57,847 (93.5%) |  |
| Yes | 2,186 (4.4%) | 541 (10.7%) | 1,274 (18.9%) | 4,001 (6.5%) |  |
| **Fever** |  |  |  |  | < 0.001 |
| No | 47,088 (94.1%) | 4,408 (87.4%) | 5,566 (82.4%) | 57,062 (92.3%) |  |
| Yes | 2,957 (5.9%) | 637 (12.6%) | 1,192 (17.6%) | 4,786 (7.7%) |  |
| **Weight loss** |  |  |  |  | < 0.001 |
| No | 49,173 (98.3%) | 4,921 (97.5%) | 6,212 (91.9%) | 60,306 (97.5%) |  |
| Yes | 872 (1.7%) | 124 (2.5%) | 546 (8.1%) | 1,542 (2.5%) |  |
| **Night sweats** |  |  |  |  | < 0.001 |
| No | 45,546 (91.0%) | 4,046 (80.2%) | 5,077 (75.1%) | 54,669 (88.4%) |  |
| Yes | 4499 (9.0%) | 999 (19.8%) | 1,681 (24.9%) | 7,179 (11.6%) |  |
| **Current TB treatment** |  |  |  |  | < 0.001 |
| No | 50,023 (100.0%) | 5,043 (100.0%) | 6,724 (99.5%) | 61,790 (99.9%) |  |
| Yes | 22 (0.0%) | 2 (0.0%) | 34 (0.5%) | 58 (0.1%) |  |
| **Previous TB treatment** |  |  |  |  | < 0.001 |
| No | 49,171 (98.3%) | 4,883 (96.8%) | 5,710 (84.5%) | 59,764 (96.6%) |  |
| Yes | 874 (1.7%) | 162 (3.2%) | 1,048 (15.5%) | 2,084 (3.4%) |  |
| **CAD4TBv6 score** |  |  |  |  | < 0.001 |
| Count | 50,045 | 5,045 | 6,758 | 61,848 |  |
| Median | 43.0 | 48.0 | 52.0 | 44.0 |  |
| Q1, Q3 | 24.0, 46.0 | 45.0, 53.0 | 46.0, 62.0 | 27.0, 48.0 |  |

\*HIV testing was not undertaken in prevalence survey. Number show participants who had their self-reported HIV status collected. <sup>†</sup>P-values calculated by Kruskal-Wallis test for CAD4TBv6 scores, Fisher’s exact test for current TB treatment, and Chi-square test for all other categorical variables.

Clinical Officers classified 50,045 (80.9%) CXRs as “normal”, 5,045 (8.2%) as “abnormal, other”, and 6,758 (10.9%) as “suggestive of TB” (Table 1). Compared to participants with CXRs classified by Clinical Officers as “normal” or “abnormal, other,” participants with CXRs classified as “suggestive of TB” were more likely to be men, self-report positive HIV status, report TB symptoms including cough, prolonged cough, fever, weight loss and night sweats, and have been previously treated for TB. CAD4TBv6 scores were significantly higher among participants whose CXR were classified as “suggestive of TB” (median: 52, IQR: 46-62), compared to those classified as “normal” (median: 43, IQR: 24-46) or “abnormal, other” (median: 48, IQR: 45-53), p<0.0001.

### CAD4TBv6 scores by sputum testing and bacteriological TB status

Sputum was collected from 8,996/61,848 (14.5%) participants, of whom 298 (3.3%) had bacteriologically-confirmed TB. CAD4TBv6 scores were higher among participants who had sputum tested in the prevalence survey (median: 49, 95%HDI: 9-82) than where sputum was not tested (median: 44, 95%HDI: 4-55) - Figure 2. The median CAD4TBv6 score for participants with bacteriologically -confirmed TB was substantially higher at 72 (95%HDI: 38-98) compared to 49 (95%HDI: 8-79) for participants with bacteriologically-negative sputum results.

**Figure 2:**
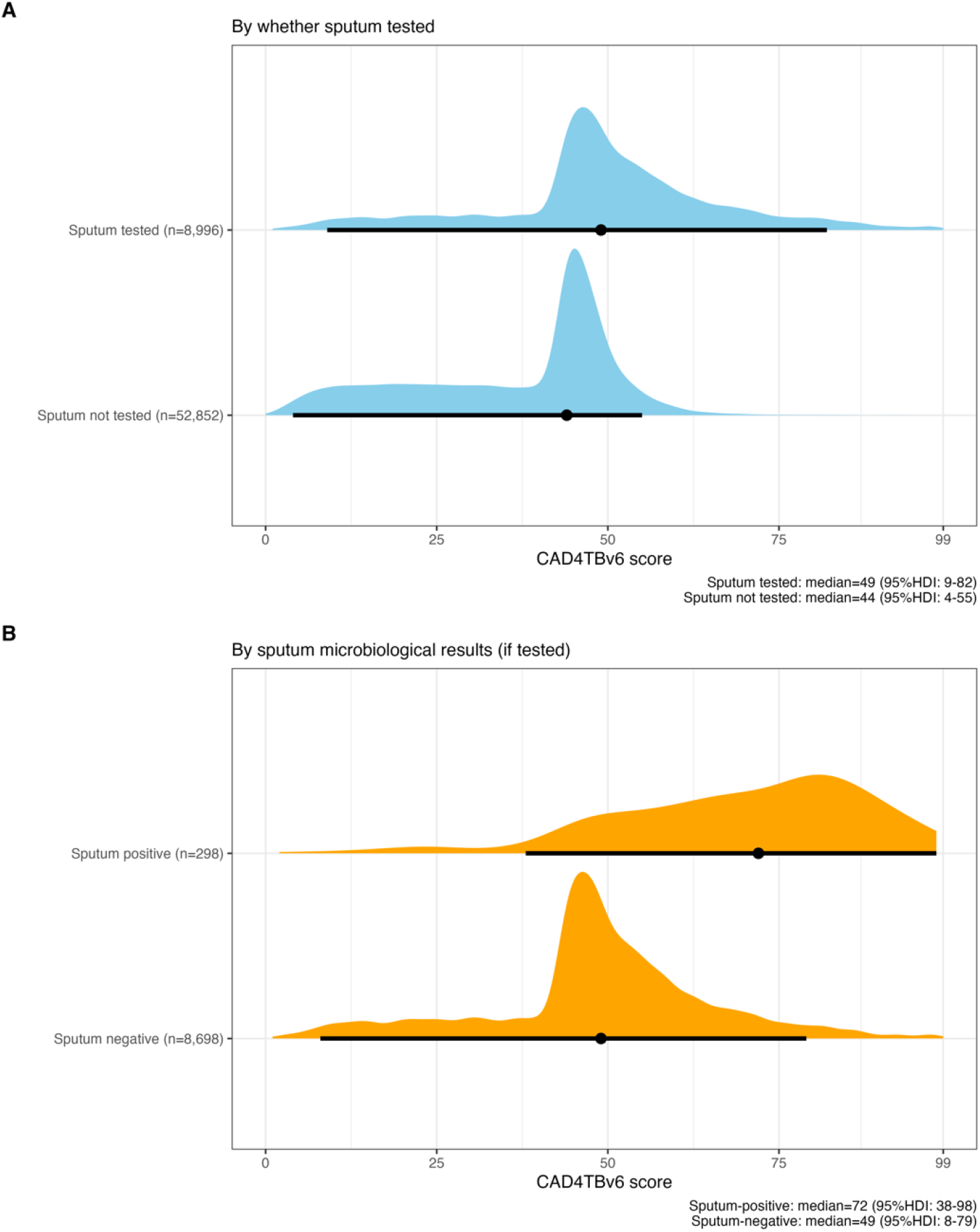
Distribution of CAD4TBv6 scores in Kenya National TB prevalence survey. A) Distribution (median and 95% highest density interval) of CAD4TBv6 scores by whether prevalence survey participant’s sputum was tested or not. B) Distribution (median and 95% highest density interval) of CAD4TBv6 scores by sputum bacteriological results. 95%HDI: 95% highest density interval.

Overall, 4,678/8,996 (52.0%) of participants with sputum collected were women and the median age was 45 years (IQR: 30-61). Among those tested, more men had bacteriologically-confirmed TB (185/4318, 4.3%) than women (113/4678, 2.4%, p<0.0001). Conditional on being tested, bacteriologically-confirmed TB was higher among those with cough of more than two weeks (3.9%, 140/3636) compared to those with no cough or cough of less than two weeks (2.9%, 158/5360, p=0.022), and among participants with weight loss (5.8%, 39/671) compared to those without weight loss (3.1%, 259/8325, p<0.0003). Conditional on testing, participants who had previous TB treatment were more likely to be bacteriologically positive (7.3%, 81/1109) than those who had not been previously treatment (2.8%, 217/7887, p<0.0001).

### Model estimated TB prevalence and accuracy of Clinical Officer CXR interpretation, sputum testing and CAD4TBv6 screening

Model Gelman-Rubin statistics were all <1.01 and trace plots showed good mixing across chains (Supplemental Text 2). Mean posterior estimates of the prevalence of bacteriologically-confirmed pulmonary TB were 568 per 100,000 (95% credible interval [CrI]: 478-654) overall, 927 per 100,000 (95% CrI: 783-1090) for men, and 314 per 100,000 (95% CrI: 246-391) for women.

From the model, we estimated that the overall sensitivity of Clinical Officer CXR interpretation as “suggestive of TB” for bacteriologically-confirmed TB was 43.7% (95% CrI: 23.8-66.4%) and specificity was 89.2% (89.0%-89.6%) – Table 2. However, in stratified analysis, sensitivity of Clinical Officer CXR interpretation was considerably higher (84.8%, 95% CrI: 70.0-94.2%) and specificity was lower (54.1% 95% CrI: 51.3-56.9%) among participants who had previously been treated for TB. Accuracy was similar when stratified by sex and chronic cough status. The posterior mean sensitivity of the combined sputum or Xpert or culture reference standard for the true underlying TB status was estimated to be 70.0% (95% CrI: 56.8%-84.6%) overall, and was 97.0% (95% CrI: 96.5%-97.5%) among participants with cough for two weeks or longer.

**Table 2:** Model-based accuracy of screening and diagnostic tests for TB latent class.

|  | Sensitivity (%) | 95% CrI | Specificity (%) | 95% CrI |
| --- | --- | --- | --- | --- |
| **Clinical Officer CXR interpretation** |  |  |  |  |
| **“suggestive of TB”** |  |  |  |  |
| Overall | 43.7% | 23.8 - 66.4% | 89.2% | 89.0 - 89.6% |
| Men | 48.4% | 27.7 - 71.0% | 87.1% | 86.6 - 87.5% |
| Women | 40.3% | 21.0 - 63.2% | 90.9% | 90.6 - 91.2% |
| Cough >2weeks | 53.5% | 33.1 - 74.2% | 83.1% | 82.7 - 83.6% |
| No cough >2weeks | 43.0% | 23.1 - 65.9% | 89.7% | 89.5 - 90.0% |
| Previous TB | 84.8% | 70.0 - 94.2% | 54.1% | 51.3 - 56.9% |
| No previous TB | 42.2% | 22.1 - 65.4% | 90.5% | 90.3 - 90.8% |
| **Sputum Xpert or culture** |  |  |  |  |
| No cough >2weeks | 70.0% | 56.8 - 84.6% | 99% (fixed) | -- |
| Cough >2weeks | 97.0% | 96.5 - 97.5% | 99% (fixed) | -- |
| **Against minimum Target Product Profile** |  |  |  |  |
| CAD4TBv6 (for sensitivity 90%) CAD: 61 | 90% (fixed) | -- | 90.4% | 88.1 - 92.6% |
| CAD4TBv6 (for specificity 70%) CAD: 47 | 98.3% | 97.6 - 98.8% | 70% (fixed) | -- |
| **Against optimum Target Product Profile** |  |  |  |  |
| CAD4TBv6 (for sensitivity 95%) CAD: 55 | 95% (fixed) | -- | 83.2% | 79.9 - 86.6% |
| CAD4TBv6 (for specificity 80%) CAD: 53 | 96.2% | 95.0 - 97.3% | 80% (fixed) | -- |

CrI: Credible interval

The posterior mean estimate of the AUC for CAD4TBv6 was 96.7% (95% CrI: 95.9%-97.4%). With CAD4TBv6 thresholds set to achieve a sensitivity of 90% (minimum TPP) or 95% (optimum TPP) mean specificity was 90.4% (95% CrI: 88.1%-92.6%) and 83.2% (95% CrI: 79.9%-86.6%) respectively –Figure 3. With CAD4TBv6 thresholds set to achieve a specificity of 70% (minimum TPP) or 80% (optimum TPP), mean sensitivity was 98.3% (95% CrI: 97.6-99.8%) and 96.2% (95.0-97.3%).

**Figure 3:**
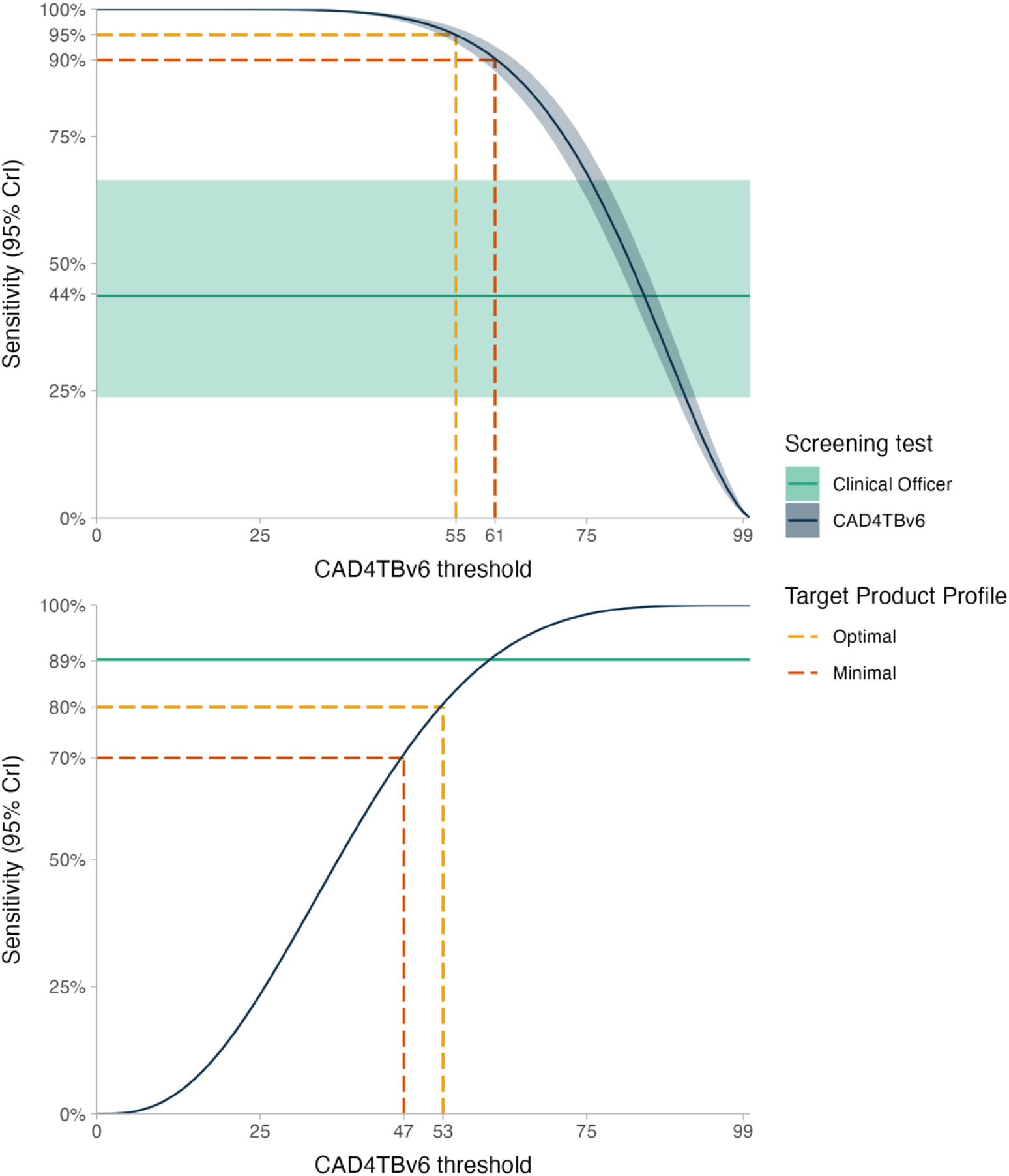
Model-based sensitivity and specificity of CAD4TBv6 for bacteriologically-confirmed pulmonary TB at minimum and optimum target product profile thresholds.

When model estimates were stratified by participant characteristics (age, sex, presence of cough of more than two weeks, history of previous TB), we found substantial variation in the sensitivity and specificity of CAD4TBv6 for bacteriologically-confirmed pulmonary TB (Figure 4). With the CAD4TBv6 threshold set to 55 to achieve overall sensitivity of 95% for the optimal TPP within the prevalence survey population, sensitivity was highest among participants aged 41 years and older, who had previously been treated for TB and who had cough for more than two weeks. In contrast, specificity was lowest among participants previously treated for TB, and among older participants.

**Figure 4:**
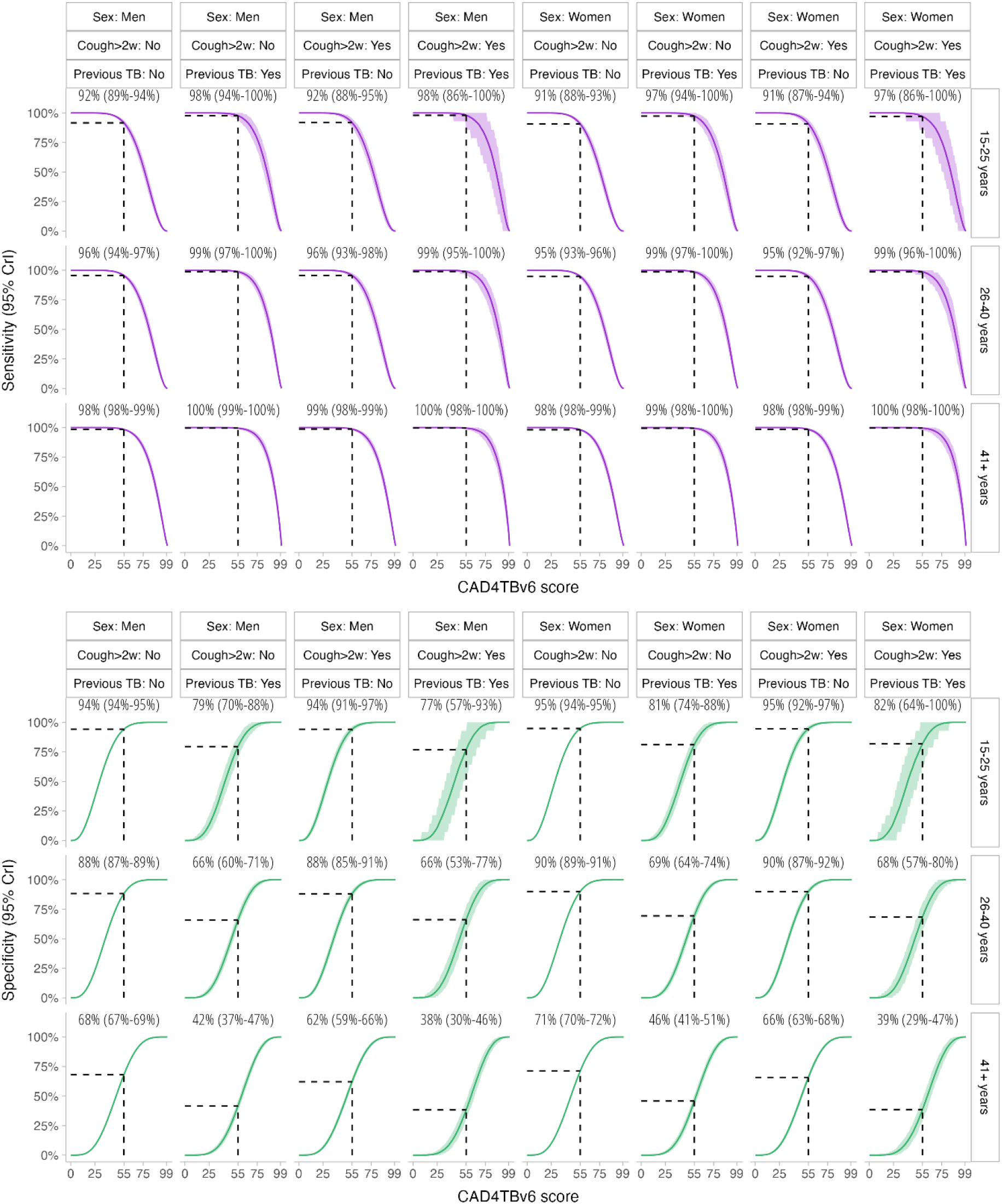
Sensitivity and specificity of CAD4TBv6 by prevalence survey participant characteristics, with threshold set at optimal target product profile to achieve overall sensitivity of 95% (CAD4TBv6=55)

## Discussion

This is the first study, to the best of our knowledge, to evaluate the accuracy of computer-aided CXR screening for TB in a community-based prevalence survey. Highly specific Xpert and culture tests were used as the bacteriological reference standard, with Bayesian latent class modelling employed to infer disease prevalence within the portion of the study population without TB symptoms or CXR signs suggestive of TB. Overall in the screening population, CAD4TBv6 met both the minimum and optimum TPP for a community-based referral test for identifying people suspected of having TB.(20) Very high sensitivity was demonstrated in participants in older age groups (41 years or older), those with reported cough>2 weeks and participants with previous TB history. Conversely, participants in older age groups and those with previous TB history had lower specificity. Computer-aided CXR screening is an accurate tool that could be used to support community TB screening in high burden countries where access to radiologists and clinicians is limited. To optimise screening accuracy and efficiency of confirmatory sputum testing, we recommend that an adaptive approach to screening threshold definition is adopted based on participant characteristics.

Community-based active case finding (ACF) for TB is effective at reducing TB prevalence if delivered with sufficient and sustained intensity to high burden populations.(25, 26) However, operationalisation of ACF in a resource limited setting has been challenging due to substantial resourcing requirements and suboptimal TB screening and diagnosis tests.(12, 27) The availability of portable/ultra-portable CXRs and CAD offer a potential solution to conduct community-based ACF for at risk groups in densely populated urban areas where TB transmission is now concentrated.(16) We have demonstrated that, overall within the prevalence survey population, CXR based screening in combination with CAD is highly accurate. CAD gives the additional flexibility for TB programs to vary the threshold for sputum testing with a saving of up to 50% of Xpert tests.(16) Given the limited resources available to National TB Programmes, by varying the CAD screening threshold, the number of TB cases deemed acceptable to be missed can be balanced against how much money is available to spend on expensive confirmatory sputum investigation.(16) By adopting an adaptive threshold within population groups, we believe that further gains in accuracy and programme efficiency can be gained. CXR and CAD as tools for community-based TB screening ACF, additionally offer the potential for individuals and TB programmes, including: earlier diagnosis; identification of asymptomatic TB, potentially reducing transmission; reductions in false positive bacteriological tests with harm from prolonged unnecessary treatment; and reduction in catastrophic costs.(13, 16, 28)

The prevalence survey participants are representative of the general population as they were randomly selected with a high participation rate, though higher amongst women than men.(18, 21) The CAD accuracy finding in our study is therefore likely to be generalizable to countries in sub-Saharan Africa with high burden of TB and HIV. Though our study focused on one software (CAD4TBv.6), other comparable software that had the CE (Conformité Européenne) marking by January 2020 (Lunit Insight CXR, Lunit Insight; and qXR v2, Qure.ai.) may perform similarly or better than this.(5, 16) Rapid advances have been made in CAD software development, with a total of 12 software solutions identified in March 2021 and version updates occurring frequently. (16, 29) Regular updating of WHO guidelines is therefore required to keep pace with these advances. As national TB programs adopt CAD technology into screening activities, in addition to performance, other implementation considerations include; cost effectiveness, compatibility of the X-ray systems, input image format, integration with any patient archiving systems, customer service and support, data protection, and ability to detect other non TB conditions.(16, 29, 30) Conditions other than TB may be as, or more prevalent than TB in high TB prevalence settings, and require comprehensive approaches to ensure participants in TB screening programmes are linked to appropriate care. (31) TB screening programs should plan for this and take into consideration resource implications to ensure additional health benefits through the identification of populations at risk of diseases other than TB. In addition to diagnostic accuracy; clinical utility, acceptability and feasibility of using CAD should be assessed.(32)

In our secondary analysis we found that accuracy varied considerably by participant characteristics, specifically age and previous TB history. Similar to a previous study in Bangladesh among adults attending primary health care for triage setting, there was no significant difference in performance of CAD4TBv6 between men and women.(16) The lower specificity of CAD4TBv6 in the older age groups and those with prior history of TB is a finding similar to previous studies.(6, 16, 17) There are numerous anatomical and pathophysiological changes occurring in old age that could explain this lower age-related specificity, including age-related changes and sequalae of life-course accumulated lung damage.(33) People with prior TB have lung changes that could lead to difficulty distinguishing old vs active disease, leading to low specificity.(6, 16, 17) Further algorithm training with images from older populations may result in refinement of CAD software with improvements in specificity. In addition, two stage screening of CAD with symptom screen followed by C-reactive protein or other novel screening tests in older populations and in participants with previous TB history could improve specificity, although this requires further investigation.

In prevalence surveys, image classification criteria are set to a low threshold for referral for sputum testing, and non-expert readers like Clinical Officers are trained to interpret with higher sensitivity and lower specificity to avoid missing prevalent TB cases.(21) We found that, overall, the sensitivity of Clinical Officer CXR interpretation (“suggestive of TB”) for the underlying true TB status was lower (44%) and specificity was higher (89%) than anticipated,(5) but that sensitivity was substantially higher (84%) and specificity lower (54%) among participants with previous TB, and with no appreciable differences by other participant characteristics. This overall low sensitivity is not usually identified by other analyses that compare clinical CXR interpretation to a microbiological reference standard, and that assume that sputum testing is 100% sensitive. From our latent class model, we can then infer that true TB cases that are bacteriologically-negative are likely to have minimal or no CXR abnormalities (unless previously treated for TB), and so are currently undetectable without new, more sensitive TB diagnostic tests. As in other studies, we have demonstrated that CAD at varied thresholds achieves higher sensitivity than human readers.(7, 14, 16) CAD has a high throughput and has been shown to reduce the time to treatment (28, 30). Additionally, CAD has the benefit of flexibility of varying thresholds, with a higher threshold improving the positive predictive value and reducing the number of Xpert tests required to diagnose a patient by up to 50% while maintaining sensitivity above 90% (16). For TB prevalence surveys, we recommend that based on accuracy, a strategy including CAD should be considered, supported by formal health economic analyses to determine the health system feasibility of wide scale implementation. Mathematical modelling studies will likely be required to investigate potential impact on longer term trends of TB incidence, prevalence and mortality.

A major strength of our study is the use of a large population based data set from a well-conducted, WHO-approved TB prevalence survey.(18, 21) Analysis of the CXRs was blinded to bacteriological status, and we used a robust bacteriological reference standard.(34) Our model prevalence estimates are slightly higher than empirical estimates obtained from the prevalence survey (especially for men: 927 per 100,000 [783-1090] in our analysis vs. 809 per 100,000 [656-962] in the weighted prevalence survey estimate) ;(18) this is to be anticipated as our model accounted for less than perfect sensitivity of sputum diagnostic tests (Xpert and culture) to estimate the underlying true prevalence of disease. Limitations include our results being model-based and are dependent on the validity of the model assumptions; to achieve computational efficiency, we fixed the specificity of the bacteriological reference standard to be 99%. Study participants were aged 15-years and above and we therefore cannot comment on the diagnostic accuracy of CAD4TBv6 in children. The study only included bacteriologically-confirmed TB; assessment of accuracy in participants with sputum negative TB (clinically diagnosed) or extra-pulmonary TB (pleural) is challenging to undertake. We also were not able to stratify performance by HIV status as testing was not systematically conducted during the prevalence survey. This would have been important for an in-depth subgroup analysis in a high TB-HIV prevalence setting. Kenya has a HIV prevalence of 4.9% with approximately 1.6 million people living with HIV and an estimated HIV-positive TB incidence at 70/100,000.(1, 35) We therefore expected a lower CAD specificity in our setting as CXR is known to be less sensitive in immunocompromised patients with pulmonary TB.(36-39) We recommend further evaluation of CAD software in high TB-HIV prevalence settings and further studies on accuracy within HIV-positive populations.

In conclusion, the END TB strategy calls for concerted efforts to improve diagnosis of TB, including through new and effective approaches to systematic screening. We have demonstrated that CAD4TBv6 is an accurate tool for community based TB screening in Kenya and met the TPP in this population. In resource limited settings where radiologists are scarce, an adaptive approach to setting screening thresholds could further improve screening accuracy and efficiency.

## Supporting information

S2: Modelling CAD4TBv6 accuracy

## Data Availability

The Kenya National Tuberculosis, Leprosy and Lung Disease program is the custodian of the 2016 Kenya Tuberculosis Prevalence Survey data.

## Contributions

PM, BNM, BM JO, EM and SBS were responsible for the study conceptualization. BNM, JO, SBS, EJ and PM designed the study. EM, JO and JS as key investigators of the Kenya prevalence survey, contributed to the study protocol development. Approval for use of the data for this study was given by JS, JO, EM and EO. BNM, EJ, BM, JO and EM developed the study protocol. BNM, SBS PM, CCK, MYRH, and JO developed study methodology. DK, RK and BNM uploaded the images for analysis. DK and RK managed the study data. PM, CCK, and MYRH conducted the data analysis and development of the figures. BNM, EJ, BM, SBS and PM played a major role in data interpretation, the writing and review of the manuscript. EM, VM, BM, B Mugi, CCK, and MYRH critically reviewed the manuscript. SBS was Director of the IMPALA Global Health Research Unit at the time of this work and played a major role in securing the funding for the work. All authors provided final approval for publication. The IMPALA consortium as a whole provided review and feedback on the progress of the work at consortium meetings or through study related advisory panels.

## Declaration of interest

We declare no competing interests.

## Acknowledgements

We are grateful to the 2016 Kenya Tuberculosis Prevalence Survey team and the Division of National Tuberculosis, Leprosy and Lung Disease Program whose data we used for this secondary study. We would like to thank Martin Githiomi-Information Technology specialist who helped in review of the DELFT-NTLD-AFIDEP contracts ensuring the data protection act was adhered to. We also acknowledge Wendy Nkirote who played a role in review of the methodology part of the prevalence survey laboratory processes to ensure it was captured accurately. Finally Maureen Kamene (Former Head NTLD Program), Aiban Ronoh (Head of department, Monitoring and Evaluation) and Jeremiah Okari (Head of department-Diagnostics) who facilitated the process of accessing the prevalence survey data.

## Funding

This research was funded by the National Institute for Health Research (NIHR) (IMPALA, grant reference 16/136/35) using UK aid from the UK Government to support global health research. The views expressed in this publication are those of the author(s) and not necessarily those of the NIHR or the UK Department of Health and Social Care. PM is funded by Wellcome (200901/Z/16/Z). MYRH was in part supported by a strategic award from Wellcome to Malawi-Liverpool-Wellcome Trust Clinical Research programme (206545/Z/17/Z).

## Supplementary material

**S1:** Prevalence Survey Clinical officers qualifications and work experience.

|  | EDUCATION QUALIFICATION | WORK EXPERIENCE |
| --- | --- | --- |
| 1 | DIPLOMA IN CLINICAL MEDICINE AND SURGERY | 4 YEARS |
| 2 | DIPLOMA IN CLINICAL MEDICINE AND SURGERY | 2 YEARS |
| 3 | DIPLOMA IN CLINICAL MEDICINE AND SURGERY AND COMMUNITY HEALTH | 1 YEAR |
| 4 | DIPLOMA IN CLINICAL MEDICINE SURGERY AND COMMUNITY HEALTH | 1 YEAR |
| 5 | DIPLOMA IN CLINICAL MEDICINE SURGERY AND COMMUNITY HEALTH | 2 YEARS |
| 6 | DIPLOMA IN CLINICAL MEDICINE AND SURGERY | 1 YEAR |
| 7 | DIPLOMA IN CLINICAL MEDICINE AND SURGERY | 1 YEAR |
| 8 | BSc ENVIRONMENTAL HEALTH, AND DIPLOMA IN CLINICAL MEDICINE AND SURGERY | 2 YEARS |
| 9 | DIPLOMA IN CLINICAL MEDICINE AND SURGERY | 2 YEARS |
| 10 | DIPLOMA IN CLINICAL MEDICINE AND SURGERY | 2 YEARS |

**S2: Modelling CAD4TBv6 accuracy**

## Notes

### Competing Interest Statement

The authors have declared no competing interest.

### Author Declarations

This study was conducted as part of the Kenya Prevalence survey ethics approval reference number SSC 2094 by the Kenya Medical Research Institute.

## References

1. World Health Organisation. Global tuberculosis report 2020. Geneva; 2020. Contract No.: ISBN 978-92-4-001313-1.

2. WHO. Tuberculosis key facts 2020 [Available from: https://www.who.int/news-room/fact-sheets/detail/tuberculosis.

3. WHO. The END TB Strategy. Geneva: World Health Organisation; 2015.

4. Chakaya J, Khan M, Ntoumi F, Aklillu E, Fatima R, Mwaba P, et al. Global Tuberculosis Report 2020 – Reflections on the Global TB burden, treatment and prevention efforts. International Journal of Infectious Diseases. 2021.

5. WHO. WHO consolidated guidelines on tuberculosis. Module 2: screening – systematic screening for tuberculosis disease. Geneva: World Health Organization 2021.

6. Qin Z, Ahmed S, Sarker MS, Paul K, Adel ASS, Naheyan T, et al. Can artificial intelligence (AI) be used to accurately detect tuberculosis (TB) from chest x-ray? A multiplatform evaluation of five AI products used for TB screening in a high TB-burden setting. ArXiv. 2020;abs/2006.05509.

7. Murphy K, Habib SS, Zaidi SMA, Khowaja S, Khan A, Melendez J, et al. Computer aided detection of tuberculosis on chest radiographs: An evaluation of the CAD4TB v6 system. Scientific Reports. 2020;10(1):5492.

8. Harris M, Qi A, Jeagal L, Torabi N, Menzies D, Korobitsyn A, et al. A systematic review of the diagnostic accuracy of artificial intelligence-based computer programs to analyze chest x-rays for pulmonary tuberculosis. PLOS ONE. 2019;14(9):e0221339.

9. Golub JE, Mohan CI, Comstock GW, Chaisson RE. Active case finding of tuberculosis: historical perspective and future prospects. Int J Tuberc Lung Dis. 2005;9(11):1183–203.

10. WHO. Systematic screening for active tuberculosis Principles and recommendations. Geneva, Switzerland: WHO Document Production Services; 2013.

11. WHO. Toman’s Tuberculosis Case detection, treatment, and monitoring – questions and answers. 2nd Edition ed. Frieden T, editor. China2004.

12. Miller C, Lonnroth K, Sotgiu G, Migliori GB. The long and winding road of chest radiography for tuberculosis detection. European Respiratory Journal. 2017;49(5):1700364.

13. Frascella B, Richards AS, Sossen B, Emery JC, Odone A, Law I, et al. Subclinical tuberculosis disease - a review and analysis of prevalence surveys to inform definitions, burden, associations and screening methodology. Clin Infect Dis. 2020.

14. Breuninger M, van Ginneken B, Philipsen RH, Mhimbira F, Hella JJ, Lwilla F, et al. Diagnostic accuracy of computer-aided detection of pulmonary tuberculosis in chest radiographs: a validation study from sub-Saharan Africa. PLoS One. 2014;9(9):e106381.

15. Qin ZZ, Sander M, Rai B, Titahong C, Sudrungrot S, Laah S, et al. Using artificial intelligence to read chest radiographs for tuberculosis detection: A multi-site evaluation of the diagnostic accuracy of three deep learning systems. Scientific Reports. 2019;9:15000.

16. Qin ZZ, Ahmed S, Sarker MS, Paul K, Adel ASS, Naheyan T, et al. Tuberculosis detection from chest x-rays for triaging in a high tuberculosis-burden setting: an evaluation of five artificial intelligence algorithms. The Lancet Digital Health. 2021;3(9):e543–e54.

17. Khan FA, Majidulla A, Tavaziva G, Nazish A, Abidi SK, Benedetti A, et al. Chest x-ray analysis with deep learning-based software as a triage test for pulmonary tuberculosis: a prospective study of diagnostic accuracy for culture-confirmed disease. The Lancet Digital Health. 2020;2(11):e573–e81.

18. Enos M, Sitienei J, Ong’ang’o J, Mungai B, Kamene M, Wambugu J, et al. Kenya tuberculosis prevalence survey 2016: Challenges and opportunities of ending TB in Kenya. PLOS ONE. 2018;13(12):e0209098.

19. NTLD. Kenya Tuberculosis Prevalence Survey 2016. Nairobi, Kenya: National Tuberculosis, Leprosy and Lung Disease Program; 2018.

20. WHO. WHO. High-priority target product profiles for new tuberculosis diagnostics: report of a consensus meeting. Geneva, Switzerland: World Health Organisation; 2014. Contract No.: WHO/HTM/TB/2014.18.

21. WHO. Tuberculosis PREVALENCE SURVEYS: a handbook. China2011.

22. Systems DI. Xray systems 2019 [Available from: https://www.delft.care/x-ray-systems/.

23. Kenya Go. DATA, SYSTEM GOVERNANCE AND CHANGE MANAGEMENT FRAMEWORK. In: Division of Health Informatics MaE, editor. Nairobi, Kenya: Ministry Of Health; 2015.

24. General Data Protection Regulation, (2016).

25. Burke RM, Nliwasa M, Feasey HRA, Chaisson LH, Golub JE, Naufal F, et al. Community-based active case-finding interventions for tuberculosis: a systematic review. The Lancet Public Health. 2021;6(5):e283–e99.

26. Marks GB, Nguyen NV, Nguyen PTB, Nguyen T-A, Nguyen HB, Tran KH, et al. Community-wide Screening for Tuberculosis in a High-Prevalence Setting. New England Journal of Medicine. 2019;381(14):1347–57.

27. Wilson JMG, Jungner G, World Health O. Principles and practice of screening for disease / J.M.G. Wilson, G. Jungner. Geneva: World Health Organization; 1968.

28. MacPherson P, Webb EL, Kamchedzera W, Joekes E, Mjoli G, Lalloo DG, et al. Computer-aided X-ray screening for tuberculosis and HIV testing among adults with cough in Malawi (the PROSPECT study): A randomised trial and cost-effectiveness analysis. PLoS Med. 2021;18(9):e1003752.

29. Stop T, Partnership. Screening and Triage for TB using Computer-Aided Detection (CAD) Technology and Ultra-portable X-Ray Systems: A Practical Guide. 2021.

30. FIND. Digital Chest Radiography and Computer-Aided Detection (CAD) Solutions for Tuberculosis Diagnostics, Technology Landscape Analysis. FIND; 2021.

31. Mungai BN, Joekes E, Masini E, Obasi A, Manduku V, Mugi B, et al. ‘If not TB, what could it be?’ Chest X-ray findings from the 2016 Kenya Tuberculosis Prevalence Survey. Thorax. 2021.

32. Cocco P, Ayaz-Shah A, Messenger MP, West RM, Shinkins B. Target Product Profiles for medical tests: a systematic review of current methods. BMC Medicine. 2020;18(1):119.

33. Hochhegger B, Meireles G, Irion K, Zanetti G, Garcia E, Moreira J, et al. The chest and aging: radiological findings. Jornal brasileiro de pneumologia : publicacao oficial da Sociedade Brasileira de Pneumologia e Tisilogia. 2012;38:656–65.

34. WHO. Xpert MTB/RIF implementation manual Technical and operational ‘how-to’: practical considerations. 2014. Contract No.: WHO/HTM/TB/2014.1.

35. (NASCOP) NAaSCP. Preliminary KENPHIA 2018 Report. Nairobi; 2020.

36. van Cleeff MR, Kivihya-Ndugga LE, Meme H, Odhiambo JA, Klatser PR. The role and performance of chest X-ray for the diagnosis of tuberculosis: a cost-effectiveness analysis in Nairobi, Kenya. BMC Infect Dis. 2005;5:111.

37. Perlman DC, el-Sadr WM, Nelson ET, Matts JP, Telzak EE, Salomon N, et al. Variation of chest radiographic patterns in pulmonary tuberculosis by degree of human immunodeficiency virus-related immunosuppression. The Terry Beirn Community Programs for Clinical Research on AIDS (CPCRA). The AIDS Clinical Trials Group (ACTG). Clin Infect Dis. 1997;25(2):242–6.

38. Philipsen RHHM, Sánchez CI, Maduskar P, Melendez J, Peters-Bax L, Peter JG, et al. Automated chest-radiography as a triage for Xpert testing in resource-constrained settings: a prospective study of diagnostic accuracy and costs. Scientific Reports. 2015;5(1):12215.

39. Muyoyeta M, Maduskar P, Moyo M, Kasese N, Milimo D, Spooner R, et al. The sensitivity and specificity of using a computer aided diagnosis program for automatically scoring chest X-rays of presumptive TB patients compared with Xpert MTB/RIF in Lusaka Zambia. PLoS One. 2014;9(4):e93757.

