## Supplementary material for "Accuracy of computer-aided chest X-ray screening in the Kenya National Tuberculosis Prevalence Survey": S2: Modelling CAD4TBv6 accuracy: 2021-09-10_analysis_kcxr (5).html

Supplemental Text 2: Analysis of CAD4TBv6 in Kenya National TB Prevalence Survey


### Supplemental Text 2: Analysis of CAD4TBv6 in Kenya National TB Prevalence Survey

This Supplemental file accompanies the manuscript: “**Accuracy of computer-aided chest X-ray screening in the Kenya National Tuberculosis Prevalence Survey**” by Mungai et al.

The Kenya National Tuberculosis, Leprosy and Lung Disease program is the custodian of the 2016 Kenya Tuberculosis Prevalence Survey data. As data cannot be made openly available, here we provide code and output to summarise the analysis and modelling approach taken.

Analysis was undertaken using R version 4.1.1.

##### 1. Packages used in analysis

```
library(tidyverse)
library(rstan)
library(tidybayes)
library(here)
library(posterior)
library(bayesplot)
library(cmdstanr)
library(arsenal)
library(scales)
library(RColorBrewer)
library(patchwork)
library(DiagrammeR)
```

##### 2. Descriptive analysis

Summarise the characteristics of the TB Prevalence Survey participants

```
#Load the raw dataset
load("kenyacad.rda")

#Some recoding and tidying up
#recode characteristics
kenyacad <- kenyacad %>%
  mutate(field_read2 =case_when(xfindingall==1 ~ "Normal",
                                xfindingall==2 ~ "Suggestive of TB",
                                xfindingall==3 ~ "Abnormal, other")) %>%
  mutate(c2w = case_when(
    coughing=="Yes" & coughingweeksnumber >=2 ~ "Cough >=2w",
    TRUE ~ "No cough >=2w"))%>%
  mutate(tbeverbeentreated = case_when(tbeverbeentreated==1 ~ "Previous_TB",
                                       tbeverbeentreated==0 ~ "Never_TB")) %>%
  mutate(hiv = case_when(hiv==1 ~ "HIV_positive",
                         hiv==0 ~ "HIV_negative")) %>%
  mutate(tbcurrenttreatment = case_when(tbcurrenttreatment=="Not taking TB treatment" ~ "Not_taking_TB_treatment",
                                        tbcurrenttreatment=="Taking TB treatment" ~ "Taking_TB_treatment")) %>%
  mutate(ageg2 = case_when(agegp=="15 - 24" ~ "below_45",
                           agegp=="25 - 34" ~ "below_45",
                           agegp=="35 - 44" ~ "below_45",
                           agegp=="45 - 54" ~ "above_45",
                           agegp=="55 - 64" ~ "above_45",
                           agegp=="65+" ~ "above_45")) %>%
  mutate(tested = case_when(xpert_culture=="Positive" | xpert_culture=="Negative" ~ "Sputum_tested",
                            is.na(xpert_culture) ~ "Sputum_not_tested"))
```

###### Table 1: characteristics of participants

```
#Set up table 1
mycontrols  <- tableby.control(
  numeric.stats=c("N", "median", "q1q3"),
  stats.labels=list(N='Count', median='Median', q1q3='Q1,Q3'),
  cat.test = "fe",
  simulate.p.value=TRUE,
  digits = 1,
  numeric.simplify = TRUE)

#rename some variables for nice printing
t1_data <- kenyacad %>%
  select(field_read2, `Age group`=agegp, Sex=sex, `HIV-status`=hiv,
         Cough=coughing, `Cough>2 weeks`=c2w, Fever=fever, `Weight loss`=weightloss,
         `Night sweats`=nightsweats, `Current TB treatment`=tbcurrenttreatment, 
         `Previous TB treatment`=tbeverbeentreated, CAD4TBv6=cad4tb6) %>%
  mutate(Sex = case_when(Sex=="M" ~ "Male",
                         Sex=="F" ~ "Female"),
         `HIV-status`=case_when(`HIV-status`=="HIV_positive" ~ "HIV-positive",
                                `HIV-status`=="HIV_negative" ~ "HIV-negative"),
         `Cough>2 weeks`=case_when(`Cough>2 weeks`=="Cough >=2w" ~ "Yes",
                                   `Cough>2 weeks`=="No cough >=2w" ~ "No"),
         `Current TB treatment` = case_when(`Current TB treatment`=="Taking_TB_treatment" ~ "Yes",
                                            `Current TB treatment`=="Not_taking_TB_treatment" ~ "No"),
         `Previous TB treatment` = case_when(`Previous TB treatment`=="Previous_TB" ~ "Yes",
                                             `Previous TB treatment`=="Never_TB" ~ "No"))

#Display table 1
t1a <- tableby(field_read2 ~ `Age group` + Sex + `HIV-status` + Cough + `Cough>2 weeks` + 
               Fever + `Weight loss` + `Night sweats` + `Current TB treatment` + 
               `Previous TB treatment` + CAD4TBv6, data=t1_data, control=mycontrols) %>%
  summary(term.name = "", text = NULL)

t1a
```

|  | Abnormal, other (N=5045) | Normal (N=50045) | Suggestive of TB (N=6758) | Total (N=61848) | p value |
| --- | --- | --- | --- | --- | --- |
| Age group |  |  |  |  | < 0.001 |
| 15 - 24 | 403 (8.0%) | 16305 (32.6%) | 742 (11.0%) | 17450 (28.2%) |  |
| 25 - 34 | 627 (12.4%) | 13400 (26.8%) | 1167 (17.3%) | 15194 (24.6%) |  |
| 35 - 44 | 765 (15.2%) | 9072 (18.1%) | 1273 (18.8%) | 11110 (18.0%) |  |
| 45 - 54 | 831 (16.5%) | 5553 (11.1%) | 1032 (15.3%) | 7416 (12.0%) |  |
| 55 - 64 | 901 (17.9%) | 3210 (6.4%) | 957 (14.2%) | 5068 (8.2%) |  |
| 65+ | 1518 (30.1%) | 2505 (5.0%) | 1587 (23.5%) | 5610 (9.1%) |  |
| Sex |  |  |  |  | < 0.001 |
| Female | 3410 (67.6%) | 29432 (58.8%) | 3345 (49.5%) | 36187 (58.5%) |  |
| Male | 1635 (32.4%) | 20613 (41.2%) | 3413 (50.5%) | 25661 (41.5%) |  |
| HIV-status |  |  |  |  | < 0.001 |
| N-Miss | 2871 | 23895 | 3587 | 30353 |  |
| HIV-negative | 2055 (94.5%) | 24985 (95.5%) | 2878 (90.8%) | 29918 (95.0%) |  |
| HIV-positive | 119 (5.5%) | 1165 (4.5%) | 293 (9.2%) | 1577 (5.0%) |  |
| Cough |  |  |  |  | < 0.001 |
| No | 4065 (80.6%) | 43989 (87.9%) | 4752 (70.3%) | 52806 (85.4%) |  |
| Yes | 980 (19.4%) | 6056 (12.1%) | 2006 (29.7%) | 9042 (14.6%) |  |
| Cough>2 weeks |  |  |  |  | < 0.001 |
| No | 4504 (89.3%) | 47859 (95.6%) | 5484 (81.1%) | 57847 (93.5%) |  |
| Yes | 541 (10.7%) | 2186 (4.4%) | 1274 (18.9%) | 4001 (6.5%) |  |
| Fever |  |  |  |  | < 0.001 |
| No | 4408 (87.4%) | 47088 (94.1%) | 5566 (82.4%) | 57062 (92.3%) |  |
| Yes | 637 (12.6%) | 2957 (5.9%) | 1192 (17.6%) | 4786 (7.7%) |  |
| Weight loss |  |  |  |  | < 0.001 |
| No | 4921 (97.5%) | 49173 (98.3%) | 6212 (91.9%) | 60306 (97.5%) |  |
| Yes | 124 (2.5%) | 872 (1.7%) | 546 (8.1%) | 1542 (2.5%) |  |
| Night sweats |  |  |  |  | < 0.001 |
| No | 4046 (80.2%) | 45546 (91.0%) | 5077 (75.1%) | 54669 (88.4%) |  |
| Yes | 999 (19.8%) | 4499 (9.0%) | 1681 (24.9%) | 7179 (11.6%) |  |
| Current TB treatment |  |  |  |  | < 0.001 |
| No | 5043 (100.0%) | 50023 (100.0%) | 6724 (99.5%) | 61790 (99.9%) |  |
| Yes | 2 (0.0%) | 22 (0.0%) | 34 (0.5%) | 58 (0.1%) |  |
| Previous TB treatment |  |  |  |  | < 0.001 |
| No | 4883 (96.8%) | 49171 (98.3%) | 5710 (84.5%) | 59764 (96.6%) |  |
| Yes | 162 (3.2%) | 874 (1.7%) | 1048 (15.5%) | 2084 (3.4%) |  |
| CAD4TBv6 |  |  |  |  | < 0.001 |
| Count | 5045 | 50045 | 6758 | 61848 |  |
| Median | 48.0 | 43.0 | 52.0 | 44.0 |  |
| Q1,Q3 | 45.0, 53.0 | 24.0, 46.0 | 46.0, 62.0 | 27.0, 48.0 |  |

###### Distribution of CAD4TBv6 scores (Figure 2).

```
s1 <- kenyacad %>%
  group_by(tested) %>%
  median_hdci(cad4tb6) %>%
  mutate(text = paste0("Median: ", cad4tb6, " (95%HDI: ",.lower, "-",.upper,")"))

s1
```

```
s2 <- kenyacad %>%
  filter(!is.na(xpert_culture)) %>%
  group_by(xpert_culture) %>%
  median_hdci(cad4tb6) %>%
  mutate(text = paste0("Median: ", cad4tb6, " (95%HDI: ",.lower, "-",.upper,")"))

s2
```

```
nrow_tested <- kenyacad %>%
  filter(tested=="Sputum_tested") %>%
  nrow(.)

nrow_nottested <- kenyacad %>%
  filter(tested=="Sputum_not_tested") %>%
  nrow(.)

p1 <- kenyacad %>% 
  mutate(tested = case_when(tested=="Sputum_tested" ~ paste0("Sputum tested ", "(n=", comma(nrow_tested),")", sep=""),
                            tested=="Sputum_not_tested" ~ paste0("Sputum not tested ", "(n=", comma(nrow_nottested),")", sep=""))) %>%
  ggplot(aes(x=cad4tb6, tested)) +
  stat_halfeye(point_interval = median_hdi, .width = c(.95), fill="skyblue") +
  theme_bw(base_size = 10) +
  scale_x_continuous(limits = c(0,99), breaks = c(0,25,50,75,99), labels = c(0,25,50,75,99)) +
  labs(x = "CAD4TBv6 score",
       y="",
       subtitle = "By whether sputum tested",
       caption = "Sputum tested: median=49 (95%HDI: 9-82)\nSputum not tested: median=44 (95%HDI: 4-55)")

p1
```

```
nrow_positive <- kenyacad %>%
  filter(xpert_culture=="Positive") %>%
  nrow(.)

nrow_negative <- kenyacad %>%
  filter(xpert_culture=="Negative") %>%
  nrow(.)

p2 <- kenyacad %>% 
  filter(!is.na(xpert_culture)) %>%
  mutate(xpert_culture = case_when(xpert_culture=="Positive" ~ paste0("Sputum positive ", "(n=", comma(nrow_positive),")", sep=""),
                                   xpert_culture=="Negative" ~ paste0("Sputum negative ", "(n=", comma(nrow_negative),")", sep=""))) %>%
  ggplot(aes(x=cad4tb6, xpert_culture)) +
  stat_halfeye(point_interval = median_hdi, .width = c(.95), fill="orange") +
  theme_bw(base_size = 10) +
  scale_x_continuous(limits = c(0,99), breaks = c(0,25,50,75,99), labels = c(0,25,50,75,99)) +
  labs(x = "CAD4TBv6 score",
       y="",
       subtitle = "By sputum microbiological results (if tested)",
       caption = "Sputum-positive: median=72 (95%HDI: 38-98)\nSputum-negative: median=49 (95%HDI: 8-79)")

p2
```

```
pw <- p1 / p2

pw + plot_annotation(
  tag_levels = 'A' )&
    theme(plot.tag = element_text(face = 'bold'),
          title = element_text(size=10))
```

```
ggsave(here("fig2.png"), width=8, height=10, dpi=600)
```

##### 3. Modelling CAD4TBv6 accuracy

The model is composed of three component models: TB prevalence, Standard testing, and CAD4TBv6 models. To compare the accuracy of standard testing and CAD4TBv6, we used a Bayesian latent class model for canonical correlation analysis for linking to TB status, which is not directly observable, instead of assuming either of them as a reference standard, to the results of the two sets of tests.

For implementing the two testing components, we used a mixture distribution approach. That is, there were two counterfactuals of TB and non-TB for each subject surveyed; each counterfactual has a respective likelihood for each test, and each subject has a probability of having TB. The likelihood of the screening models is the average likelihood of test results for TB and non-TB weighted by the probabilities of having TB or not. Across all tests included, we applied the assumption that we assumed both standard testing and CAD4TBv6 are better than flipping a fair coin, at least. That is, the TB status relates a higher chance of positivity from standard TB screening and a higher CAD4TBv6 score.

###### TB status model (modelling TB prevalence by the covariates)

The TB prevalence model links the determinants of prevalent TB and the TB status. We structure the TB prevalence model with a logistic regression model for estimating the TB prevalence. The TB status is not directly supported by the data, so the probability of TB is related to the test results of standard testing and CAD4TBv6. The sub-model is formulated as follows:

\[\begin{align}
D\_i &\sim Bernouilli(p\_i)\\
p\_i &= logit(p\_0) + \lambda X \\
logit(p\_0) &\sim Normal(-5.219282, 0.4076352) \\
\lambda &\sim Normal(0, 1)
\end{align}\]

where \(d\_i\) denotes the status of TB disease of subject \(i\) which is a latent class variable, \(\lambda\) denotes the coefficients linking the covariates of \(X\) to the probability of \(D\_i\). The prior for \(logit(p\_0)\) was inferred from the results of the TB prevalence survey.

###### CAD4TBv6

We modelled CAD4TBv6 with Beta regression for proportional data or other data bounded between 0 and 1. The CAD4TB evaluates CXR images and scores them from 0 to 99. We remap the scores to a variable ranging from 0 to 1.

The CAD4TBv6 model is formulated as follows:

For the counterfactual with TB (\(D\_i = 1\)):

\[\begin{align}
Y\_{CAD, i} &\sim Beta(gA\_1, gB\_1)\\
gA\_1 &= \nu\_1 \kappa \\
gB\_1 &= (1 - \nu\_1) \kappa \\
logit(\nu\_1) &= \gamma\_1 + \gamma X
\end{align}\]

For the counterfactual without TB (\(d\_i = 0\)):

\[\begin{align}
Y\_{CAD, i} &\sim Beta(gA\_0, gB\_0)\\
gA\_0 &= \nu\_0 \kappa \\
gB\_0 &= (1 - \nu\_0) \kappa \\
logit(\nu\_0) &= \gamma\_0 + \gamma X
\end{align}\]

with prior distributions: \[\begin{align}
\gamma &\sim Normal(0, 10) \\
\gamma\_0 &\sim Normal(0, 10) \\
\gamma\_1 - \gamma\_0 &\sim HalfNormal(0, 1) \\
\kappa &\sim inverseGamma(1, 1)
\end{align}\]

where \(Y\_{CAD, i}\) denotes the score of CAD4TBv6,;`\(\gamma\) denotes the coefficients linking the covariates of \(X\), \(\gamma\_1\) and \(\gamma\_0\) are the intercept terms of the regression function with and without TB respectively; \(\kappa\) is a scale parameter determining the variance of the distribution. Lastly, we weighted the two counterfactuals by the probabilities of \(D\_i = 1\) or not.

###### Standard testing

In this study, standard testing is a TB detection approach used commonly in TB prevalence surveys. The algorithm started with a CXR read by a human reader and initial consultation followed by a sputum microbiological testing on those with CXR-abnormalities or TB-suggestive symptoms (here cough >=2 weeks).

We modelled the results of CXR read by human readers with a logistic regression model. The TB and non-TB models share the same coefficients of predictors, while the counterfactual with TB has a larger value of intercept term which implied the subjects with TB are more likely to have positive results.

Following CXR read by human readers, we modelled the results of sputum microbiological testing with a Bernoulli process with the probability of sensitivity of TB and (1 – specificity) of non-TB. Considering the quality of sputum samples are symptom related, we assumed the sensitivity differs by coughing >= 2 weeks status. For an identifiability issue that the sensitivity and the specificity cannot simultaneously be characterised in the model, we used an exogenously given specificity of 99%.

The standard testing model is formulated as follows:

For the counterfactual with TB (\(D\_i = 1\)):

\[\begin{align}
Y\_{CO, i} &\sim Bernouilli(r\_i)\\
r\_i &= \beta\_0 + \beta X\\
\beta &\sim Normal(0, 10) \\
\beta\_0 &\sim Normal(0, 10)
\end{align}\] if sputum sample collected and tested: \[\begin{align}
Y\_{Sput, i} &\sim Bernouilli(sens\_{sput})\\
\end{align}\]

For the counterfactual without TB (\(D\_i = 0\)):

\[\begin{align}
Y\_{CO, i} &\sim Bernouilli(r\_i)\\
r\_i &= \beta\_1 + \beta X\\
\end{align}\] if sputum sample collected and tested: \[\begin{align}
Y\_{Sput, i} &\sim Bernouilli(1 - spec\_{sput})
\end{align}\]

with prior distributions: \[\begin{align}
\beta &\sim Normal(0, 10) \\
\beta\_0 &\sim Normal(0, 10) \\
\beta\_1 - \beta\_0 &\sim HalfNormal(0, 1) \\
2\*(sens\_{sput} - 0.5) &\sim Beta(2, 4)
\end{align}\]

where \(Y\_{CO, i}\) is the results of CXR read by clinical officers; \(\beta\) denotes the coefficients linking the covariates of \(X\) to CXR positivities; \(\beta\_1\) and \(\beta\_0\) are the intercept terms of the regression function with and without TB respectively. \(Y\_{Sput, i}\) is the results of sputum tests; \(sens\_{sput}\) and \(spec\_{sput}\) denote the sensitivity and specificity of sputum tests the values related with/without coughing \(\ge\) 2 weeks. Lastly, we weighted the two counterfactuals by the probabilities of \(D\_i = 1\) or not.

###### Modelling running, diagnostics and outputs

Code below runs the Stan model (is computationally intensive, so commented out here).

```
##read the tidied dataset
# dat <- read_rds("2021-06-17_kcxr.rds")

# #Define model inputs as a list
# inputs <- with(dat, {
#   list(
#     N = nrow(dat),
#     CAD = CAD,
#     CO = CO,
#     Spu = Spu,
#     SpuMis = c(SpuMis),
#     N_Cov = 3,
#     Covs = cbind(Male, Before, Age_centre),
#     Cough2w = c(Cough2w),
#     spec_sput = 0.99,
#     spec_sput_c2w = 0.99
#   )
# })

# #Define modelling sampling parameters
# n_iter = 4000 #Number of posterior samples
# n_collect = 2000 #Number of post warm-up samples
# n_chain = 3 #number of chains
# n_core = 1 #number of cores 

## Set up the model
## This calls the model_c_mono.stan file
# file <- file.path("model_c_mono.stan")


##Now fit the model
##WARNING: THIS IS SLOW AND MEMORY INTENSIVE!!!
##Saves about a 30Gb object
# model <- cmdstan_model(file)

##Now extract the posterios from the cmdstanr model object
# csv_files <- c("model_c_mono-202110040818-1-03568b.csv",
#                "model_c_mono-202110040818-2-03568b.csv",
#                "model_c_mono-202110040818-3-03568b.csv")
# 
# x <- read_cmdstan_csv(
#     csv_files,
#     variables = c("sens_sput", "sens_sput_c2w", "lambda", "beta", "gamma", "gamma0", "gamma1", "k", "beta0", "beta1", "logit_p"),
#     sampler_diagnostics = ""
# )
# 
# saveRDS(x, "x.rds")

# 
# csv_files <- c("model_c_mono-202110040818-1-03568b.csv",
#                "model_c_mono-202110040818-2-03568b.csv",
#                "model_c_mono-202110040818-3-03568b.csv")
# 
# co_out <- read_cmdstan_csv(
#     csv_files,
#     variables = c("sens_co", "spec_co"),
#     sampler_diagnostics = ""
# )
# 
# saveRDS(co_out, "co_out.rds")
```

Read in the data and the modelling output.

```
#read in the cmdstanr model output object
posterior <- read_rds(here("posterior.rds"))
#read the data in
dat <- read_rds("2021-06-17_kcxr.rds")
dat <- dat %>%
  mutate(Age_centre = scale(Age))

#read in the posterior samples
x <- read_rds(here("x.rds"))
co_out <- read_rds("co_out.rds")


#The inputs that we used for the model.
#Needed for the summarise function
inputs <- with(dat, {
  list(
    N = nrow(dat),
    CAD = CAD,
    CO = CO,
    Spu = Spu,
    SpuMis = c(SpuMis),
    N_Cov = 3,
    Covs = cbind(Male, Before, Age_centre),
    Cough2w = c(Cough2w),
    spec_sput = 0.99,
    spec_sput_c2w = 0.99
  )
})
```

###### Modelling summary

```
posterior::summarise_draws(x$post_warmup_draws)
```

```
posterior::summarise_draws(co_out$post_warmup_draws)
```

```
mcmc_trace(x$post_warmup_draws)
```

```
mcmc_trace(co_out$post_warmup_draws)
```

###### Model-based prevalence estimates

Function to summarise prevalence estimates.

```
summarise_prev <- function(post, inputs, keys, dat, itermax = 1000) {
  require(tidyverse)
  require(posterior)
  inv_logit <- function(x) { exp(x) / (1 + exp(x))}
  
  covs <- inputs$Covs
  
  pars <- as_tibble(as_draws_df(post$post_warmup_draws))
  
  pars <- list(
    lambda = pars %>% select(starts_with("lambda[")) %>% as.matrix(),
    logit_p = pars %>% select(logit_p) %>% pull()
  )
  
  n_iter <- nrow(pars$gamma)
  n_sample <- nrow(covs)
  
  itermax <- min(n_iter, itermax)
  
  # calculate prevalence
  prev <- bind_rows(lapply(1:itermax, function(i) {
    tibble(ID = keys, MC = i,
           p = inv_logit(pars$logit_p[i] + colSums(t(covs) * pars$lambda[i, ])))
  })) %>% filter(ID %in% keys)
  
  
  prev %>% left_join(dat %>% mutate(ID = 1:n()))
}


#Overall prevalence
selected <- 1:inputs$N # all samples
prev <- summarise_prev(x, inputs, keys=selected, dat)
```

```
## Joining, by = "ID"
```

```
Summary_Prev <- bind_rows(
  prev %>% 
    group_by(MC) %>% 
    summarise(p = sum(p) / n()) %>% 
    ungroup() %>% 
    summarise(
      Group = "All",
      Prev = mean(p),
      Prev_L = quantile(p, 0.025),
      Prev_U = quantile(p, 0.975),
    ),
  prev %>% 
    group_by(MC, Male) %>% ## select MC and grouping
    summarise(p = sum(p) / n()) %>% 
    group_by(Male) %>% ## keep grouping only
    summarise(
      Group="Stratified",
      Prev = mean(p),
      Prev_L = quantile(p, 0.025),
      Prev_U = quantile(p, 0.975),
    ) %>% ungroup()
)
```

```
## `summarise()` has grouped output by 'MC'. You can override using the `.groups` argument.
```

```
Summary_Prev %>%
  mutate_at(.vars = vars(Prev, Prev_L, Prev_U),
            .funs = funs(.*100000))
```

```
## Warning: `funs()` was deprecated in dplyr 0.8.0.
## Please use a list of either functions or lambdas: 
## 
##   # Simple named list: 
##   list(mean = mean, median = median)
## 
##   # Auto named with `tibble::lst()`: 
##   tibble::lst(mean, median)
## 
##   # Using lambdas
##   list(~ mean(., trim = .2), ~ median(., na.rm = TRUE))
## This warning is displayed once every 8 hours.
## Call `lifecycle::last_lifecycle_warnings()` to see where this warning was generated.
```

Clinical Officer read accuracy

```
summarise_co <- function(posterior, inputs, keys, itermax = 1000) {
  require(tidyverse)
  require(posterior)

  inv_logit <- function(x) { exp(x) / (1 + exp(x))}
  logit <- function(p) {log(p / (1 - p))}

  covs <- inputs$Covs

  pars <- as_tibble(as_draws_df(posterior$post_warmup_draws))

  pars <- list(
    beta0 = pars %>% select(beta0) %>% pull(),
    beta1 = pars %>% select(beta1) %>% pull(),
    beta = pars %>% select(starts_with("beta[")) %>% as.matrix(),
    k = pars %>% select(k) %>% pull()
  )


  n_iter <- nrow(pars$beta)
  n_sample <- nrow(covs)

  if (missing(keys)) keys <- 1:n_sample

  itermax <- min(n_iter, itermax)


  covs <- covs[keys, ]


  # sample clinical officer values
  co <- bind_rows(lapply(1:itermax, function(i) {
    tibble(ID = keys, MC = i,
           p0 = inv_logit(pars$beta0[i] + colSums(t(covs) * pars$beta[i, ])),
           p1 = inv_logit(pars$beta1[i] + colSums(t(covs) * pars$beta[i, ])))
  })) %>% filter(ID %in% keys) %>%
    group_by(MC) %>%
    summarise(
      SensCO = mean(p1),
      SpecCO = mean(1 - p0)
    ) %>%
    ungroup() %>%
    summarise(
      across(c(SensCO, SpecCO), list(
        M = mean,
        L = function(x) quantile(x, 0.025),
        U = function(x) quantile(x, 0.975)
      ))
    ) %>%
    pivot_longer(everything(), names_to = "Index") %>%
    tidyr::extract(Index, c("Index", "Stat"), "(\\w+)_(M|L|U)") %>%
    pivot_wider(names_from = Stat)

  co
}

itermax <- 1000

sel_all <- 1:inputs$N
sel_new <- which(inputs$Covs[, 2] == 0)
sel_rel <- which(inputs$Covs[, 2] == 1)

co_all <- summarise_co(posterior = x, inputs = inputs, keys = sel_all, itermax = itermax)
co_new <- summarise_co(posterior = x, inputs = inputs, keys = sel_new, itermax = itermax)
co_rel <- summarise_co(posterior = x, inputs = inputs, keys = sel_rel, itermax = itermax)

co_all
```

```
co_new
```

```
co_rel
```

The accuracy of Clinical Officer then appears to vary substantially by previous TB status, with sensitivity being much higher in those with a history of previous TB treatment, and lower in those not previously treated for TB.

```
sel_female <- which(inputs$Covs[, 1] == 0)
sel_male <- which(inputs$Covs[, 1] == 1)

co_male <- summarise_co(posterior = x, inputs = inputs, keys = sel_male, itermax = itermax)
co_female <- summarise_co(posterior = x, inputs = inputs, keys = sel_female, itermax = itermax)

co_male
```

```
co_female
```

No difference by sex though.

```
sel_nocough <- which(inputs$Cough2w == 0)
sel_cough2w <- which(inputs$Cough2w == 1)

co_nocough <- summarise_co(posterior = x, inputs = inputs, keys = sel_nocough, itermax = itermax)
co_cough2w <- summarise_co(posterior = x, inputs = inputs, keys = sel_cough2w, itermax = itermax)

co_nocough
```

```
co_cough2w
```

And sensitivity was a little higher among those with cough of >2w.

Function to summarise conditional posterior draws

```
summarise_cad <- function(posterior, inputs, keys, itermax = 1000) {
  require(tidyverse)
  require(posterior)
  inv_logit <- function(x) { exp(x) / (1 + exp(x))}
  
  covs <- inputs$Covs
  
  pars <- as_tibble(as_draws_df(x$post_warmup_draws))
  
  pars <- list(
    gamma0 = pars %>% select(gamma0) %>% pull(),
    gamma1 = pars %>% select(gamma1) %>% pull(),
    gamma = pars %>% select(starts_with("gamma[")) %>% as.matrix(),
    k = pars %>% select(k) %>% pull()
  )
  
  n_iter <- nrow(pars$gamma)
  n_sample <- nrow(covs)
  
  if (missing(keys)) keys <- 1:n_sample

  itermax <- min(n_iter, itermax)
  
  
  covs <- covs[keys, ]
  
  
  # sample CAD values
  cad <- bind_rows(lapply(1:itermax, function(i) {
    tibble(ID = keys, MC = i,
           nu0 = inv_logit(pars$gamma0[i] + colSums(t(covs) * pars$gamma[i, ])),
           nu1 = inv_logit(pars$gamma1[i] + colSums(t(covs) * pars$gamma[i, ]))) %>% 
      mutate(
        gA0 = nu0 * pars$k[i],
        gB0 = (1 - nu0) * pars$k[i],
        gA1 = nu1 * pars$k[i],
        gB1 = (1 - nu1) * pars$k[i],
        CAD0 = rbeta(n(), gA0, gB0),
        CAD1 = rbeta(n(), gA1, gB1)
      )
  })) %>% filter(ID %in% keys)
  
  
  # calculate sensivity and specificity by cut points
  rocs <- bind_rows(lapply(seq(0, 1, 0.01), function(cut) {
    cad %>%
      group_by(MC) %>%
      summarise(Sens = mean(CAD1 > cut), Spec = mean(CAD0 < cut)) %>%
      mutate(Cut = cut)
  }))
  
  # ROC intervals based on sensitivity
  rocs_mci_sens <- bind_rows(lapply(unique(rocs$MC) , function(mc) {
    sel <- rocs %>%
      filter(MC == mc)
    as_tibble(approx(sel$Sens, sel$Spec, seq(0, 1, 0.01))) %>%
      rename(Sens = x, Spec = y) %>%
      mutate(MC = mc)
  })) %>%
    group_by(Sens) %>%
    summarise(Spec_M = median(Spec), Spec_L = quantile(Spec, .025), Spec_U = quantile(Spec, .975))
  
  # ROC intervals based on specificity
  rocs_mci_spec <- bind_rows(lapply(unique(rocs$MC) , function(mc) {
    sel <- rocs %>%
      filter(MC == mc)
    as_tibble(approx(sel$Spec, sel$Sens, seq(0, 1, 0.01))) %>%
      rename(Spec = x, Sens = y) %>%
      mutate(MC = mc)
  })) %>%
    group_by(Spec) %>%
    summarise(Sens_M = median(Sens), Sens_L = quantile(Sens, .025), Sens_U = quantile(Sens, .975))
  
  
  
  # Calculate area under ROC curves
  auc <- rocs %>%
    group_by(MC) %>%
    summarise(AUC = sum((Sens + lag(Sens))[-1] * diff(Spec)) / 2)
  
  auc_mci <- (auc %>% arrange(AUC))[round(quantile(1:itermax, c(0.025, 0.5, 0.975))), ]
  
  # Keep curves at median and 95% credible interval
  rocs_mci <- rocs %>%
    filter(MC %in% auc_mci$MC) %>%
    mutate(
      Index = case_when(
        MC == auc_mci$MC[1] ~ "L",
        MC == auc_mci$MC[2] ~ "M",
        MC == auc_mci$MC[3] ~ "U"
      )
    ) %>%
    arrange(MC, Cut) %>%
    select(-MC) %>%
    pivot_wider(values_from = c(Sens, Spec), names_from = Index)
  
  
  list(
    ROC = rocs,
    AUC = auc,
    ROC_mci = rocs_mci,
    ROC_mci_sens = rocs_mci_sens,
    ROC_mci_spec = rocs_mci_spec
  )
}
```

###### Summarise the posterior samples

This is slow and memory intensive, so commented out here.

```
# selected <- 1:inputs$N # all samples
# 
# ### itermax: size of posterior to be used in the calculation
# 
# CAD <- summarise_cad(posterior = x, inputs = inputs, keys = selected, itermax = itermax)
# 
# saveRDS(CAD, "CAD.rds")
```

```
#read in the summarised chains
CAD <- read_rds(here("CAD.rds"))

roc <- CAD$ROC
roc_mci_sens <- CAD$ROC_mci_sens
roc_mci_spec <- CAD$ROC_mci_spec
roc_mci <- CAD$ROC_mci
auc <- CAD$AUC

auc %>% summarise(mean_auc = mean(AUC),
                  q2.5_AUC = quantile(AUC, probs = 0.025),
                  q97.5_AUC = quantile(AUC, probs = 0.975))
```

Summarise and plot the overall sensitivity and specificity by CAD4TBv6 threshold

```
roc_mci_sens %>%
  filter(Sens >0.94 & Sens <0.96)
```

```
#CAD score = 55

roc_mci_sens %>%
  filter(Sens >0.89 & Sens<0.91)
```

```
#CAD score = 61


sens_plot <- CAD$ROC %>%
  group_by(Cut) %>%
  summarise(
    Sens_M = median(Sens),
    Sens_L = quantile(Sens, 0.025),
    Sens_U = quantile(Sens, 0.975)
  ) %>%
  ggplot() +
  
  geom_rect(data=co_all %>% filter(Index=="SensCO"),
               aes(ymin=L,ymax=U, xmin=0, xmax=100,
                   fill="#1B9E77"), alpha=0.3) +
  
  geom_hline(data= co_all %>% filter(Index=="SensCO"),
            aes(yintercept=M,
                   colour="#1B9E77")) +
  
  geom_segment(aes(y=0.95, yend=0.95,
                   x=0, xend=55, alpha=0.99), colour="#EFA00B", linetype=5)+
  
  geom_segment(aes(x=55, xend=55,
                   y=0, yend=0.95,alpha=0.99), colour="#EFA00B", linetype=5) +
  
  geom_segment(aes(y=0.90, yend=0.90,
                   x=0, xend=61, alpha=0.98), colour="#D65108", linetype=5) +
  
  geom_segment(aes(x=61, xend=61,
                   y=0, yend=0.90, alpha=0.98), colour="#D65108", linetype=5) +
  
  
  geom_ribbon(aes(x=Cut*100, ymin=Sens_L, ymax = Sens_U, fill="#0D324D"), alpha=0.3) +
  geom_line(aes(x=Cut*100, y=Sens_M, colour="#0D324D")) +
  theme_ggdist() +
  labs(x="CAD4TBv6 threshold",
       y="Sensitivity (95% CrI)") +
  scale_y_continuous(expand = c(0, 0), 
                     labels = scales::percent_format(accuracy = 1), 
                     breaks = c(0,0.25, 0.44, 0.50,0.75,0.90,0.95,1.0)) +
  scale_x_continuous(expand = c(0, 0), breaks = c(0,25,55,61,75,99)) +
  scale_fill_identity(name = "Screening test",
                          breaks = c("#1B9E77", "#0D324D"),
                          labels = c("Clinical Officer","CAD4TBv6"),
                          guide = "legend") +
  scale_colour_identity(name = "Screening test",
                          breaks = c("#1B9E77", "#0D324D"),
                          labels = c("Clinical Officer","CAD4TBv6"),
                          guide = "legend") +
  scale_alpha_identity(name = "Target Product Profile",
                          breaks = c(0.99, 0.98),
                          labels = c("Optimal","Minimal"),
                          guide=guide_legend(override.aes = 
                                               list(colour=c("#EFA00B", "#D65108"))))

sens_plot
```

Specificity

```
roc_mci_spec %>%
  filter(Spec >0.79 & Spec <0.81)
```

```
#CAD score=53

roc_mci_spec %>%
  filter(Spec >0.69 & Spec<0.71)
```

```
#CAD score = 47


spec_plot <- CAD$ROC %>%
  group_by(Cut) %>%
  summarise(
    Spec_M = median(Spec),
    Spec_L = quantile(Spec, 0.025),
    Spec_U = quantile(Spec, 0.975)
  ) %>%
  ggplot() +
    
  geom_rect(data=co_all %>% filter(Index=="SpecCO"),
               aes(ymin=L,ymax=U, xmin=0, xmax=100,
                   fill="#1B9E77"), alpha=0.3) +
  
  geom_hline(data= co_all %>% filter(Index=="SpecCO"),
            aes(yintercept=M,
                   colour="#1B9E77")) +
  
  geom_segment(aes(y=0.80, yend=0.80,
                   x=0, xend=53, alpha=0.99), colour="#EFA00B", linetype=5)+
  
  geom_segment(aes(x=53, xend=53,
                   y=0, yend=0.80,alpha=0.99), colour="#EFA00B", linetype=5) +
  
  geom_segment(aes(y=0.70, yend=0.70,
                   x=0, xend=47, alpha=0.98), colour="#D65108", linetype=5) +
  
  geom_segment(aes(x=47, xend=47,
                   y=0, yend=0.70, alpha=0.98), colour="#D65108", linetype=5) +
  
  geom_ribbon(aes(x=Cut*100, ymin=Spec_L, ymax = Spec_M, fill="#0D324D"), alpha=0.3) +
  geom_line(aes(x=Cut*100, y=Spec_M, colour="#0D324D")) +
  theme_ggdist() +
  labs(x="CAD4TBv6 threshold",
       y="Sensitivity (95% CrI)") +
  scale_y_continuous(expand = c(0, 0), 
                     labels = scales::percent_format(accuracy=1), 
                     breaks = c(0,0.25,0.50,0.70,0.80,0.89,1.0)) +
  scale_x_continuous(expand = c(0, 0), breaks = c(0,25,47,53,75,99)) +
  scale_fill_identity(name = "Screening test",
                          breaks = c("#1B9E77", "#0D324D"),
                          labels = c("Clinical Officer","CAD4TBv6"),
                          guide = "legend") +
  scale_colour_identity(name = "Screening test",
                          breaks = c("#1B9E77", "#0D324D"),
                          labels = c("Clinical Officer","CAD4TBv6"),
                          guide = "legend") +
  scale_alpha_identity(name = "Target Product Profile",
                          breaks = c(0.99, 0.98),
                          labels = c("Optimal","Minimal"),
                          guide=guide_legend(override.aes = 
                                               list(colour=c("#EFA00B", "#D65108"))))

spec_plot
```

Combine together for Figure 3.

```
f3 <- sens_plot / spec_plot

f3 + plot_layout(guides = "collect")
```

```
ggsave("fig3.png", height = 8)
```

```
## Saving 7 x 8 in image
```

###### Analysis stratified by participant characteristics

Function to summarise stratified accuracy estimates.

```
cad_selector <- function(data=dat, Male, Cough2w, Before, Age1, Age2, Age3){
  data %>%
  mutate(id=row_number()) %>%
  filter(Male=={{Male}} & Cough2w=={{Cough2w}} & Before=={{Before}} 
         & Age1=={{Age1}} & Age2=={{Age2}} & Age3=={{Age3}}) %>%
  pull(id)
}

selected <- crossing(Male=dat$Male, Cough2w=dat$Cough2w,
                     Before=dat$Before,  Age1=dat$Age1, 
                     Age2=dat$Age2, Age3=dat$Age3) %>%
  filter(Age1 + Age2 + Age3 == 1) %>%
  mutate(selected = pmap(list(Male, Cough2w, Before, Age1, Age2, Age3),
                   ~ cad_selector(Male = ..1,
                                  Cough2w = ..2,
                                  Before = ..3,
                                  Age1 = ..4,
                                  Age2 = ..5,
                                  Age3 = ..6)))

itermax <- 600


##Stratified by participant characteristics
##This is very memory intensive, so commented out here.
# CAD_stratifed <- selected %>%
#   mutate(id=1:n()) %>%
#    rowwise() %>%
#    mutate(
#    sum = list(summarise_cad(posterior = posterior, inputs = inputs, keys = selected, itermax = itermax)))
# 
# write_rds(CAD_stratifed, here("CAD_stratified.rds"))
```

Estimate stratified accuracy

```
CAD_stratified <- read_rds(here("CAD_stratified.rds"))

sums_stratified <- CAD_stratified %>%
  hoist(sum,
        ROC="ROC",
        AUC="AUC",
        ROC_mci = "ROC_mci"
        )

rocs_stratified <- sums_stratified %>%
  select(Male, Cough2w, ROC, Age1, Age2, Age3, Before) %>%
  unnest() %>%
  mutate(Test = "CAD4TB")
```

```
## Warning: `cols` is now required when using unnest().
## Please use `cols = c(ROC)`
```

```
aucs_stratified <- sums_stratified %>%
  select(Male, Cough2w, AUC, Age1, Age2, Age3, Before) %>%
  unnest()%>%
  mutate(Test = "CAD4TB")
```

```
## Warning: `cols` is now required when using unnest().
## Please use `cols = c(AUC)`
```

```
aucs_sum_stratified <- aucs_stratified %>%
  summarise(M = median(AUC), L = quantile(AUC, 0.025), U = quantile(AUC, 0.975)) %>% 
  as.list()
```

Plot the stratified estimates of accuracy (Figure 4)

```
z <- rocs_stratified %>%
  pivot_longer(c(Sens, Spec), names_to = "Index") %>%
  mutate(Cut = as.numeric(scales::number(Cut*100, accuracy = 1))) %>%
  mutate_at(.vars = vars(Male, Cough2w, Age1, Age2, Age3, Before),
            .funs = funs(case_when(
              .==0 ~ "No",
              .==1 ~ "Yes"
            ))) %>%  
  mutate(Age = case_when(Age1=="No" & Age2=="No" & Age3=="Yes" ~ "41+ years",
                         Age1=="No" & Age2=="Yes" & Age3=="No" ~ "26-40 years",
                         Age1=="Yes" & Age2=="No" & Age3=="No" ~ "15-25 years")) %>%
  mutate(Index = case_when(Index=="Sens" ~ "Sensitivity",
                           Index=="Spec" ~ "Specificity")) %>%
  mutate(Male = case_when(Male=="No" ~ "Women",
                          Male=="Yes" ~ "Men"))

z_h <- z %>%
  mutate_at(.vars = vars(Male, Cough2w,  Age, Before),
            .funs = funs(factor)) %>%
  pivot_wider(names_from = Index,
              values_from = value) 


z2 <- z %>%
  filter(Cut==55) %>%
  group_by(Male, Cough2w,  Index, Before, Age, Cut) %>% 
  summarise(mean = mean(value),
            l = quantile(value, probs=0.025),
            u = quantile(value, probs=0.975)) %>%
  mutate(posx = 90,
         posy = 0.75) %>%
  rename(Sex = Male)
```

```
## `summarise()` has grouped output by 'Male', 'Cough2w', 'Index', 'Before', 'Age'. You can override using the `.groups` argument.
```

```
z2 <- z2 %>%
  mutate(mci = paste0(scales::percent(mean, accuracy = 1), " (", scales::percent(l, accuracy = 1), "-", scales::percent(u, accuracy = 1), ")"))


sens_plot2 <- z_h %>%
  group_by(Cut, Male, Cough2w, Before, Age) %>%
    rename(`Cough>2w` = Cough2w,
         `Previous TB` = Before,
         Sex = Male) %>%
  summarise(Sens_M = mean(Sensitivity),
            Sens_L = quantile(Sensitivity, probs=0.025),
            Sens_U = quantile(Sensitivity, probs=0.975)) %>%
  ggplot() +
  geom_line(aes(x=Cut, y=Sens_M), colour="darkorchid") +
  geom_ribbon(aes(x=Cut, ymin=Sens_L, ymax=Sens_U), alpha=0.3, fill="darkorchid") +
  geom_segment(data=z2 %>% filter(Index=="Sensitivity")%>%
    rename(`Cough>2w` = Cough2w,
         `Previous TB` = Before), aes(x=0, xend=Cut, y=mean, yend=mean), linetype="dashed") +
  geom_segment(data=z2 %>% filter(Index=="Sensitivity")%>%
    rename(`Cough>2w` = Cough2w,
         `Previous TB` = Before), aes(x=Cut, xend=Cut, y=0, yend=mean), linetype="dashed") +
  geom_text(data=z2 %>% filter(Index=="Sensitivity")%>%
    rename(`Cough>2w` = Cough2w,
         `Previous TB` = Before), aes(x=50, y=1.15, label=mci),
            family="Open Sans Condensed Light") +
  scale_y_continuous("Sensitivity (95% CrI)", labels = scales::percent, breaks = c(0,0.25,0.50,0.75,1)) +
  facet_grid(Age ~ Sex + `Cough>2w` + `Previous TB`, scales = "free_x", 
             labeller = labeller(.cols = label_both)) +
  theme_ggdist()+
  scale_x_continuous("CAD4TBv6 score", breaks = c(0,25, 55, 75, 99)) +
  theme(strip.background =element_rect(fill=NA, colour="darkgrey"))
```

```
## `summarise()` has grouped output by 'Cut', 'Sex', 'Cough>2w', 'Previous TB'. You can override using the `.groups` argument.
```

```
spec_plot2 <-z_h %>%
  group_by(Cut, Male, Cough2w, Before, Age) %>%
  rename(`Cough>2w` = Cough2w,
         `Previous TB` = Before,
         Sex = Male) %>%
  summarise(Spec_M = mean(Specificity),
            Spec_L = quantile(Specificity, probs=0.025),
            Spec_U = quantile(Specificity, probs=0.975)) %>%
  ggplot() +
  geom_line(aes(x=Cut, y=Spec_M), colour="mediumseagreen") +
  geom_ribbon(aes(x=Cut, ymin=Spec_L, ymax=Spec_U), alpha=0.3, fill="mediumseagreen") +
  geom_segment(data=z2 %>% filter(Index=="Specificity")%>%
    rename(`Cough>2w` = Cough2w,
         `Previous TB` = Before), aes(x=0, xend=Cut, y=mean, yend=mean), linetype="dashed") +
  geom_segment(data=z2 %>% filter(Index=="Specificity")%>%
    rename(`Cough>2w` = Cough2w,
         `Previous TB` = Before), aes(x=Cut, xend=Cut, y=0, yend=mean), linetype="dashed") +
  geom_text(data=z2 %>% filter(Index=="Specificity") %>%
    rename(`Cough>2w` = Cough2w,
         `Previous TB` = Before), aes(x=50, y=1.15, label=mci),
            family="Open Sans Condensed Light") +
  scale_y_continuous("Specificity (95% CrI)", labels = scales::percent, breaks = c(0,0.25,0.50,0.75,1)) +
  facet_grid(Age ~ Sex + `Cough>2w` + `Previous TB`, scales = "free_x", 
             labeller = labeller(.cols = label_both)) +
  theme_ggdist()+
  scale_x_continuous("CAD4TBv6 score", breaks = c(0,25, 55, 75, 99))+
  theme(strip.background =element_rect(fill=NA, colour="darkgrey"))
```

```
## `summarise()` has grouped output by 'Cut', 'Sex', 'Cough>2w', 'Previous TB'. You can override using the `.groups` argument.
```

```
sens_plot2 / spec_plot2
```

```
ggsave("fig4.png", height=12, width=10)
```
